# Stricker Learning Span criterion validity: a remote self-administered multi-device compatible digital word list memory measure shows similar ability to differentiate amyloid and tau PET-defined biomarker groups as in-person Auditory Verbal Learning Test

**DOI:** 10.1101/2022.10.26.22281534

**Authors:** Nikki H. Stricker, John L. Stricker, Ryan D. Frank, Winnie Z. Fan, Teresa J. Christianson, Jay S. Patel, Aimee J. Karstens, Walter K. Kremers, Mary M. Machulda, Julie A. Fields, Jonathan Graff-Radford, Clifford R. Jack, David S. Knopman, Michelle M. Mielke, Ronald C. Petersen

## Abstract

**Objective:** The Stricker Learning Span (SLS) is a computer-adaptive digital word list memory test specifically designed for remote assessment and self-administration on a web-based multi-device platform (Mayo Test Drive). We aimed to establish criterion validity of the SLS by comparing its ability to differentiate biomarker-defined groups to the person-administered Rey’s Auditory Verbal Learning Test (AVLT).

**Participants and Methods:** Participants (N=353; mean age=71, SD=11; 93% cognitively unimpaired [CU]) completed the AVLT during an in-person visit, the SLS remotely (within 3 months) and had brain amyloid and tau PET scans available (within 3 years). Overlapping groups were formed for 1) those on the Alzheimer’s disease (AD) continuum (amyloid PET positive, A+, n=125) or not (A-, n=228), and those with biological AD (amyloid and tau PET positive, A+T+, n=55) vs no evidence of AD pathology (A-T-, n=195). Analyses were repeated among CU participants only.

**Results:** The SLS and AVLT showed similar ability to differentiate biomarker-defined groups when comparing AUROCs (*p*’s>.05). In logistic regression models, SLS contributed significantly to predicting biomarker group beyond age, education and sex, including when limited to CU participants. Medium (A- vs A+) to large (A-T- vs A+T+) unadjusted effect sizes were observed for both SLS and AVLT. Learning and delay variables were similar in terms of ability to separate biomarker groups.

**Conclusions:** Remotely administered SLS performed similarly to in-person-administered AVLT in its ability to separate biomarker-defined groups, providing evidence of criterion validity. Results suggest the SLS may be sensitive to detecting subtle objective cognitive decline in preclinical AD.

## 1. Introduction

The need for efficient and scalable approaches for identifying individuals at risk for preclinical and prodromal Alzheimer’s disease (PAD) is paramount to ongoing clinical trial efforts, emerging decentralized trials, and for identifying individuals who will most benefit from currently available pharmacologic or behavioral treatments, or those on the horizon^1,2^. Self-administered cognitive measures that can be completed “remotely” (i.e., outside of a typical clinical setting, including at home) are a critical component of an early PAD detection strategy^3,4^. Frequently, tests originally designed for and validated within clinic settings are converted to remote use to increase access for those unable to readily visit research centers or due to necessity during the COVID-pandemic^5-7^. The limitation of this “conversion” approach is that tests are not developed specifically with remote self-administration as a priority for test design decisions, which can contribute to mixed findings when performance is compared across settings^8-10^. There is an urgent need for valid self-administered cognitive assessment tools designed specifically for remote use. Verbal memory measures are among the most sensitive to early changes in the Alzheimer’s disease (AD) process^11^ but are also challenging to adapt to remote, self-administered methods^5^.

The Stricker Learning Span (SLS) is a digital computer-adaptive word list memory test specifically designed for remote assessment^12^. The SLS is administered via a web-based multi-device platform designed for unsupervised self-administration of digital cognitive tests, Mayo Test Drive (MTD): test Development through Rapid Iteration, Validation and Expansion (DRIVE)^13^. Recent work has highlighted learning as a key deficit in PAD, conceptualized as a failure to benefit from repeated exposure^14^ or a lack of benefit from practice^15,16^. In line with this, SLS test design emphasizes learning. The SLS paradigm was influenced by cognitive science principles and neural network process simulations^12^. The SLS stresses the contextual system during learning through use of high frequency word stimuli and variations in word item-level imagery to increase difficulty.

Preliminary support for the feasibility, reliability and validity of the SLS was previously reported in an all-female older adult sample^13^. Whereas that prior study used traditional approaches to test validation, the current study aimed to establish test validity using a novel approach to avoid the inherent issues with existing validation approaches. For example, one common approach is to correlate a new test with existing cognitive tests. However, existing tests, while well established, are themselves imperfect measures of hypothesized underlying constructs^17^. Another frequent approach is to establish validity by examining the ability of a new test to differentiate clinically defined groups. In the AD field, for example, it is common to establish the clinical validity of a new test by comparing individuals who are cognitively “normal” or unimpaired to individuals with mild cognitive impairment (MCI) or dementia; however, this introduces circularity because the use of cognitive tests is central to establishing those syndromal classifications. In vivo biomarkers offer an alternative ground truth for test validation studies that is completely independent of cognitive test performance. This is akin to validation with neuropathological diagnosis at autopsy given the correspondence between antemortem PET imaging and autopsy findings but has the notable benefit of being feasible during life^18,19^. A research framework is now available to use available AD biomarkers to characterize participants using the amyloid (A), tau (T) and neurodegeneration (N), or the AT(N), system^20^. Imaging biomarkers of N are considered non-specific to AD and will not be included in the current manuscript to limit the number of subgroups. Individuals with evidence of elevated amyloid (A+) are considered to show Alzheimer’s pathologic change. An *in vivo* biological diagnosis of AD is defined by the presence of both A+ and elevated tau (T+).

The objective of this study was to determine the criterion validity of the SLS. Critically, this validation study was limited to unsupervised completion of the SLS in a remote environment outside of a typical clinical research setting. Our primary study hypothesis (Aim 1) was that remotely administered SLS and in-person-administered Rey’s Auditory Verbal Learning Test (AVLT) would differentiate AD biomarker-defined groups similarly. This hypothesis is tested in groups defined by biomarker status alone (A+ vs A- and A+T+ vs A-T-) to avoid circularity. Secondary hypotheses included that the SLS would be sensitive to preclinical AD in analyses limited to CU participants (Aim 2), SLS and AVLT would show significant correlations to support convergent validity (Aim 3), and that word list learning vs. delay indices would show similar sensitivity to biologically defined AD (A-T- vs A+T+; Aim 4).

## 2. Methods

Most participants were recruited from the Mayo Clinic Study of Aging (MCSA), a longitudinal population-based study of aging among Olmsted County, Minnesota, residents. Participants are randomly sampled by age- and sex-stratified groups using the resources of the Rochester Epidemiology Project medical records-linkage system, which links the medical records from all county providers^21^. All participants are without dementia at MCSA enrollment. Exclusion criteria include terminal illness or hospice. Participants complete study visits every 15 months that include a physician exam, study coordinator interview, and neuropsychological testing^22^. The physician exam includes a medical history review, complete neurological exam, and the Short Test of Mental Status (STMS)^23^. The study coordinator interview with an informant includes the Clinical Dementia Rating® scale^24^. Participants complete multi-domain battery of nine neuropsychological tests administered by a psychometrist^22^. The interviewing study coordinator, examining physician, and neuropsychologist initially each make an independent diagnostic determination. A final diagnosis of cognitively unimpaired, MCI^25^ or dementia^26^ is then achieved through consensus agreement^22,25^. The diagnostic evaluation does not consider prior clinical information, prior diagnoses, SLS performance, or knowledge of biomarkers.

To enrich the sample for participants with cognitive impairment, additional participants were recruited from the Mayo Alzheimer’s Disease Research Center (ADRC).

The study protocols were approved by the Mayo Clinic and Olmsted Medical Center Institutional Review Boards. All participants provided informed consent.

### *In Vivo* Neuroimaging Markers of Amyloid and Tau

The most recent imaging available ±3 years of baseline SLS was used. Amyloid and tau positivity is determined using Pittsburgh Compound B PET (PiB-PET) and tau PET (flortaucipir)^27-29^. PET images are acquired using a GE Discovery RX or DXT PET/CT scanner. A global cortical PiB PET standard uptake value ratio (SUVR) is computed by calculating the median uptake over voxels in the prefrontal, orbitofrontal, parietal, temporal, anterior cingulate, and posterior cingulate/precuneus regions of interest (ROIs) for each participant and dividing this by the median uptake over voxels in the cerebellar crus gray matter; for tau PET we utilize median uptake over the voxels in the meta regions consisting of entorhinal, amygdala, parahippocampal, fusiform, inferior temporal, and middle temporal ROIs normalized to the cerebellar crus gray matter^28^. Cutoffs to determine amyloid and tau positivity are SUVR ≥ 1.48 (centiloid 22)^30^ and ≥ 1.25^28^ respectively, to maintain consistency with our past Cogstate-focused work^31-33^.

### Person-administered AVLT completed in clinic

The psychometrist reads a list of 15 words (List A) aloud and asks the participant to repeat back as many words as they can recall. This is repeated 5 times (learning trials 1-5 total). A distractor list (List B) is presented, followed by short-delay (Trial 6) of list A words. Recall of List A is again tested after 30-minutes (30-minute delay), followed by written recognition^34,35^. The primary variable for this study is sum of trials (trials 1–5 total + trial 6 + 30-min recall; range 0-105), which is sensitive to early changes in memory^36^. Additional variables include correct words on trials 1-5 total and trial 5 (thought to reflect learning), as well as 30-minute delay. Long-term percentage retention (AVLT 30-minute delay / Trial 5), thought to reflect delayed memory or storage/savings, is also reported.

### Self-administered SLS completed remotely (not in clinic)

All participants completed the SLS remotely and without supervision or assistance. Participants followed a link provided in an email to complete the test session. The SLS is administered via the Mayo Test Drive (MTD) platform^13^.

The SLS is a 5-trial adaptive list learning task. Single words are visually presented sequentially during learning trials. After each list presentation, memory for the word list is tested with 4-choice recognition. Following a computer adaptive testing approach, the SLS starts with 8 items and then the number of words either stays the same, increases by 5 or decreases by 2 according to pre-specified rules based on percentage of correct responses to extend the floor and ceiling relative to traditional word list memory tests (range 2-23 words; **Figure 1**). Short delay follows the Symbols Test^13,37,38^; all items presented on any learning trial are tested during delay (range 8-23).

**Figure 1.**
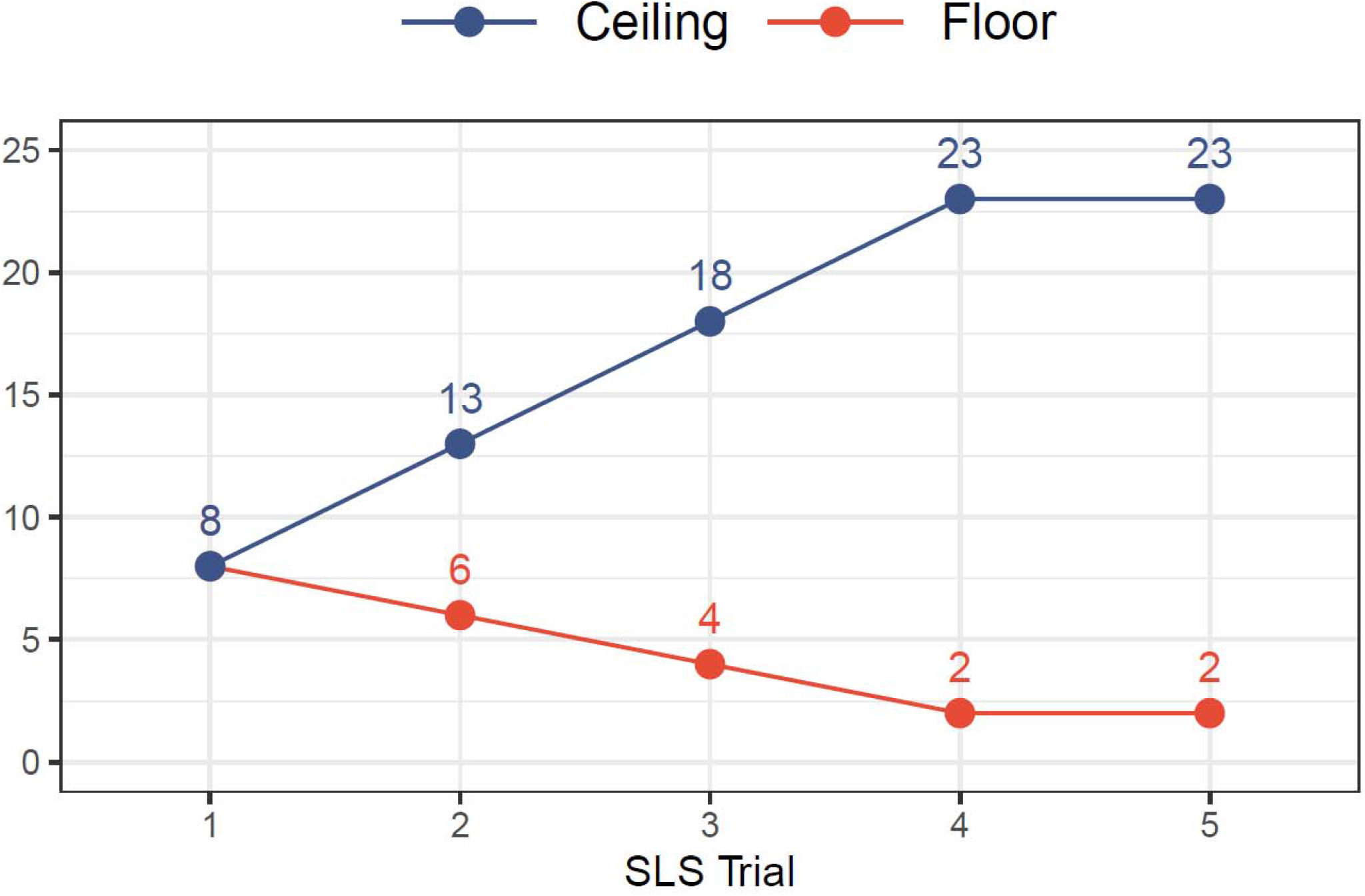
Stricker Learning Span (SLS) computer adaptive testing approach provides an expanded ceiling and floor relative to traditional word list memory tests. Figure used with permission of Mayo Foundation for Medical Education and Research; all rights reserved.

The SLS uses common, high frequency words that are easier to recall, but harder to recognize^39^, as previously described^12^. There are 23 item bins with 4 words each, and words within a bin have similar imagery ratings^40^. Each successive item bin has lower imagery ratings, thus increasing the difficulty of subsequent items. Most (90%) of the 92 total words used are on the Dolch sight words reading list (preschool through Grade 3), with half at the preschool level^41^. Each test session randomly selects 1 word from each item bin as the “target,” the 3 others serve as the foils and a 23-item target word list is generated, even if not all items are presented due to the adaptive procedure. To reduce recency effects, the order of item presentation is randomized for each trial and the last item presented is never the first tested. The primary variable is sum of trials (total correct across learning trials 1-5 plus delay, range 0-108). Secondary variables include maximum (max) learning span across any trial (range 0-23), total correct across learning trials (1-5 total, range 0-85), and total correct short delay (range 0-23). Percent retention (max span / delay) is also reported to allow within-test comparisons, but this measure is not meant to be compared to AVLT percent retention as differences are expected based on differences in test design.

### Inclusion Criteria

To be included in this study, participants had to have both SLS sum of trials and AVLT sum of trials available and an amyloid PET scan within 3 years. Participants who completed the SLS as of 7/7/22 were included in this manuscript. Available linked data as of 8/22/22 were included.

### Statistical methods

Demographics and clinical characteristics were descriptively summarized using counts and percentages for categorical data and means and standard deviations for continuous data. Data distributions across groups (A- vs A+ and A-T- vs A+T+) were compared using chi-square tests for categorical variables and linear model ANOVA tests for continuous variables. Pearson correlation coefficients were used to characterize the linear relationship between AVLT and SLS variables. Unadjusted and adjusted Hedge’s G with weighted and pooled standard deviation was used to assess effect size for group comparisons. Unadjusted and adjusted logistic regression models were used to determine the predictive accuracy of AVLT and SLS sum of trials in predicting abnormal amyloid PET (A+ vs A-) and abnormal amyloid and tau PET (A+T+ vs A-T-). To formally compare the ability of both tests to differentiate biomarker-defined groups, the AUROC from models with AVLT were directly compared to models with SLS^42^. For models that adjusted for demographic variables, age, sex, and education were the adjustment terms for both Hedge’s G and logistic regression models. A 2-sided p-value <0.05 was considered statistically significant. All analyses were performed using R version 4.1.2.

## 3. Results

### 3.1 Participant Characteristics and Convergent Validity

The mean age of the 353 participants was 71.8 (SD=10.8) years, mean education was 15.7 (SD=2.4), 53.5% were male, 98.0% were White and 92.6% were cognitively unimpaired (**Table 1**). MTD remote testing was completed within half a month (mean) of the in-person visit. Participant characteristics by biomarker subgroups are reported in **Table 2**. As expected, based on known increases in A+ and T+ rates with increased age^27-29^, biomarker positive groups were older than biomarker negative groups (*p*’s < .05). Biomarker positive and negative groups showed similar years of education and sex distribution (*p*’s > .05).

**Table 1.** Participant demographic, clinical and neuroimaging characteristics and devices used for Mayo Test Drive (mean, SD except where otherwise noted) for all participants (N=353).

|  | Mean (SD) | Range |
| --- | --- | --- |
| Age at MCSA or ADRC visit<sup>1</sup> | 70.84 (10.82) | 35-94 |
| In-person visit to MTD, months | 0.55 (0.42) | -0.30 – 2.99 |
| In-person visit to imaging, months | -0.78 (1.03) | -2.83 – 0.32 |
| Sex, N (%) Male | 189 (53.5%) | - |
| Education, years | 15.72 (2.36) | 11-20 |
| Race (% White)<sup>2</sup> | 346 (98.0%) | - |
| Ethnicity (% Non-Hispanic)<sup>3</sup> | 351 (99.7%) | - |
| Kokmen Short Test of Mental Status<sup>4</sup> | 35.6 (2.4) | 22-38 |
| N (%) first MCSA or ADRC visit | 60 (17.0%) | 1-14 |
| Diagnosis |  | - |
| Cognitively Unimpaired (N, %) | 326 (92.6%) | - |
| MCI (N, %) | 21 (6.0%) | - |
| Dementia (N, %) | 5 (1.4%) | - |
| Unavailable (N, %)<sup>5</sup> | 1 (0.3%) | - |
| CDR Global score |  | - |
| 0 (N, %) | 332 (94.1%) | - |
| 0.5 (N, %) | 20 (5.7%) | - |
| 1 (N, %) | 1 (0.3%) | - |
| Subjective memory concerns (N, % Yes)<sup>6</sup> | 201 (58.3%) | - |
| Amyloid meta-ROI | 1.54 (0.35) | 1.16 – 3.31 |
| Tau meta-ROI<sup>7</sup> | 1.21 (0.13) | 0.90 – 2.31 |
| Entorhinal Tau<sup>7</sup> | 1.13 (0.16) | 0.78 – 2.30 |
| SLS sum of trials | 74.06 (18.27) | 16-108 |
| AVLT sum of trials | 64.38 (18.77) | 17-105 |
| SLS Max Span | 16.43 (4.18) | 3-23 |
| AVLT Trial 5 | 11.06 (2.75) | 0-15 |
| SLS 1-5 Total | 59.45 (14.07) | 13-85 |
| AVLT 1-5 Total | 45.99 (12.11) | 15-75 |
| SLS Delay | 14.60 (4.61) | 3-23 |
| AVLT 30-min delay | 9.06 (3.98) | 0-15 |
| SLS Retention | 88.28 (15.11) | 36.4-133.3 |
| AVLT Retention <sup>8</sup> | 79.03 (27.80) | 0-214.3 |
| AVLT Trial 6 (short-delay) | 9.33 (3.59) | 0-15 |
| AVLT Recognition Percent Correct | 91.31 (9.34) | 53-100 |
| Mayo Test Drive device type used |  | - |
| Desktop computer or laptop (N, %) | 220 (62.3%) | - |
| Smartphone (N, %) | 95 (26.9%) | - |
| Tablet (N, %) | 37 (10.5%) | - |
| Other / Not Sure (N, %) | 1 (0.3%) | - |

*Note:* ADRC = Mayo Alzheimer's Disease Research Center; AVLT = Auditory Verbal Learning Test; AVLT Sum of Trials = AVLT 1-5 total + Trial 6 + 30-minute delay; AVLT Recognition Percent Correct = {[recognition hits+(15 – recognition false positive errors)]/30} x 100; AVLT Retention = AVLT 30-minute delay / Trial 5; CDR = Clinical Dementia Rating Scale; MCSA = Mayo Clinic Study of Aging; MTD = Mayo Test Drive; SLS = Stricker Learning Span; SLS Max Span = maximum number of words recognized across any learning trial; SLS 1-5 Total = sum of words correctly recognized across trials 1-5; SLS Retention = SLS Delay / SLS Max Span; SLS Sum of Trials = SLS 1-5 total + delay. Table used with permission of Mayo Foundation for Medical Education and Research; all rights reserved.
<sup>1</sup> n=8 Mayo Alzheimer's Disease Research Center (ADRC); n=345 Mayo Clinic Study of Aging (MCSA)
<sup>2</sup> n=2 Asian, n=3 Black, n=2 More than one
<sup>3</sup> n=1 missing
<sup>4</sup> n=1 missing
<sup>5</sup> consensus diagnosis not yet available for this participant; CDR = 0; amyloid and tau PET negative status.
<sup>6</sup> n=8 missing; subjective memory concern is "yes" when subjective memory complaints are on the Blessed Memory Test questions 1-4 is marked as worse or question 5 indicates any other problems with thinking or memory.
<sup>7</sup> n=2 missing
<sup>8</sup> n=1 missing due to AVLT Trial 5 score of 0.

**Table 2.**
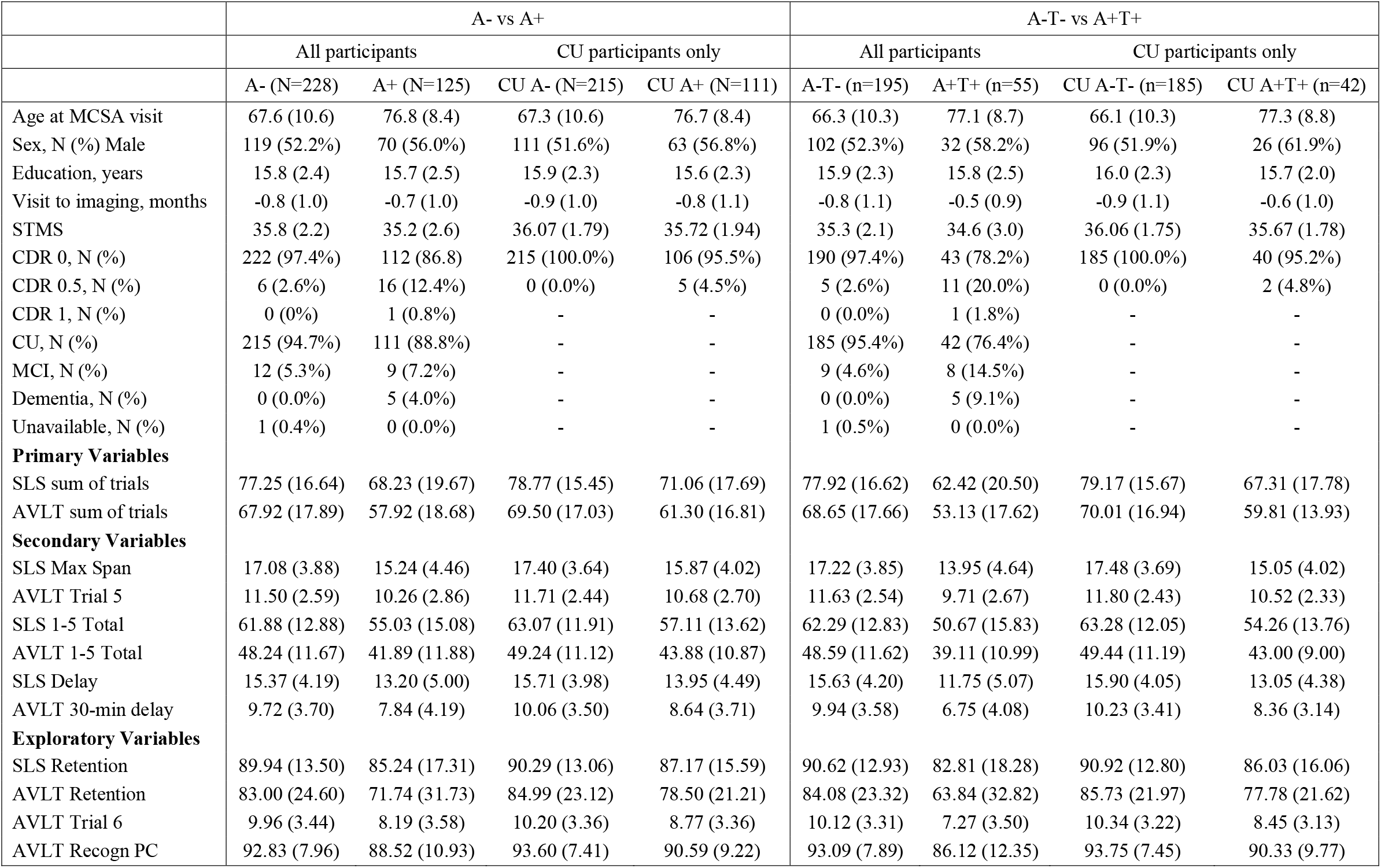
Demographic and clinical characteristics of biomarker subgroups and means (SDs) for cognitive variables.

|  | A- vs A+ |  |  |  | A-T- vs A+T+ |  |  |  |
| --- | --- | --- | --- | --- | --- | --- | --- | --- |
|  | All participants |  | CU participants only |  | All participants |  | CU participants only |  |
|  | A- (N=228) | A+ (N=125) | CU A- (N=215) | CU A+ (N=111) | A-T- (n=195) | A+T+ (n=55) | CU A-T- (n=185) | CU A+T+ (n=42) |
| Age at MCSA visit | 67.6 (10.6) | 76.8 (8.4) | 67.3 (10.6) | 76.7 (8.4) | 66.3 (10.3) | 77.1 (8.7) | 66.1 (10.3) | 77.3 (8.8) |
| Sex, N (%) Male | 119 (52.2%) | 70 (56.0%) | 111 (51.6%) | 63 (56.8%) | 102 (52.3%) | 32 (58.2%) | 96 (51.9%) | 26 (61.9%) |
| Education, years | 15.8 (2.4) | 15.7 (2.5) | 15.9 (2.3) | 15.6 (2.3) | 15.9 (2.3) | 15.8 (2.5) | 16.0 (2.3) | 15.7 (2.0) |
| Visit to imaging, months | -0.8 (1.0) | -0.7 (1.0) | -0.9 (1.0) | -0.8 (1.1) | -0.8 (1.1) | -0.5 (0.9) | -0.9 (1.1) | -0.6 (1.0) |
| STMS | 35.8 (2.2) | 35.2 (2.6) | 36.07 (1.79) | 35.72 (1.94) | 35.3 (2.1) | 34.6 (3.0) | 36.06 (1.75) | 35.67 (1.78) |
| CDR 0, N (%) | 222 (97.4%) | 112 (86.8) | 215 (100.0%) | 106 (95.5%) | 190 (97.4%) | 43 (78.2%) | 185 (100.0%) | 40 (95.2%) |
| CDR 0.5, N (%) | 6 (2.6%) | 16 (12.4%) | 0 (0.0%) | 5 (4.5%) | 5 (2.6%) | 11 (20.0%) | 0 (0.0%) | 2 (4.8%) |
| CDR 1, N (%) | 0 (0%) | 1 (0.8%) | - | - | 0 (0.0%) | 1 (1.8%) | - | - |
| CU, N (%) | 215 (94.7%) | 111 (88.8%) | - | - | 185 (95.4%) | 42 (76.4%) | - | - |
| MCI, N (%) | 12 (5.3%) | 9 (7.2%) | - | - | 9 (4.6%) | 8 (14.5%) | - | - |
| Dementia, N (%) | 0 (0.0%) | 5 (4.0%) | - | - | 0 (0.0%) | 5 (9.1%) | - | - |
| Unavailable, N (%) | 1 (0.4%) | 0 (0.0%) | - | - | 1 (0.5%) | 0 (0.0%) | - | - |
| **Primary Variables** |  |  |  |  |  |  |  |  |
| SLS sum of trials | 77.25 (16.64) | 68.23 (19.67) | 78.77 (15.45) | 71.06 (17.69) | 77.92 (16.62) | 62.42 (20.50) | 79.17 (15.67) | 67.31 (17.78) |
| AVLT sum of trials | 67.92 (17.89) | 57.92 (18.68) | 69.50 (17.03) | 61.30 (16.81) | 68.65 (17.66) | 53.13 (17.62) | 70.01 (16.94) | 59.81 (13.93) |
| **Secondary Variables** |  |  |  |  |  |  |  |  |
| SLS Max Span | 17.08 (3.88) | 15.24 (4.46) | 17.40 (3.64) | 15.87 (4.02) | 17.22 (3.85) | 13.95 (4.64) | 17.48 (3.69) | 15.05 (4.02) |
| AVLT Trial 5 | 11.50 (2.59) | 10.26 (2.86) | 11.71 (2.44) | 10.68 (2.70) | 11.63 (2.54) | 9.71 (2.67) | 11.80 (2.43) | 10.52 (2.33) |
| SLS 1-5 Total | 61.88 (12.88) | 55.03 (15.08) | 63.07 (11.91) | 57.11 (13.62) | 62.29 (12.83) | 50.67 (15.83) | 63.28 (12.05) | 54.26 (13.76) |
| AVLT 1-5 Total | 48.24 (11.67) | 41.89 (11.88) | 49.24 (11.12) | 43.88 (10.87) | 48.59 (11.62) | 39.11 (10.99) | 49.44 (11.19) | 43.00 (9.00) |
| SLS Delay | 15.37 (4.19) | 13.20 (5.00) | 15.71 (3.98) | 13.95 (4.49) | 15.63 (4.20) | 11.75 (5.07) | 15.90 (4.05) | 13.05 (4.38) |
| AVLT 30-min delay | 9.72 (3.70) | 7.84 (4.19) | 10.06 (3.50) | 8.64 (3.71) | 9.94 (3.58) | 6.75 (4.08) | 10.23 (3.41) | 8.36 (3.14) |
| **Exploratory Variables** |  |  |  |  |  |  |  |  |
| SLS Retention | 89.94 (13.50) | 85.24 (17.31) | 90.29 (13.06) | 87.17 (15.59) | 90.62 (12.93) | 82.81 (18.28) | 90.92 (12.80) | 86.03 (16.06) |
| AVLT Retention | 83.00 (24.60) | 71.74 (31.73) | 84.99 (23.12) | 78.50 (21.21) | 84.08 (23.32) | 63.84 (32.82) | 85.73 (21.97) | 77.78 (21.62) |
| AVLT Trial 6 | 9.96 (3.44) | 8.19 (3.58) | 10.20 (3.36) | 8.77 (3.36) | 10.12 (3.31) | 7.27 (3.50) | 10.34 (3.22) | 8.45 (3.13) |
| AVLT Recogn PC | 92.83 (7.96) | 88.52 (10.93) | 93.60 (7.41) | 90.59 (9.22) | 93.09 (7.89) | 86.12 (12.35) | 93.75 (7.45) | 90.33 (9.77) |

SLS sum of trials and AVLT sum of trials were strongly correlated (r = 0.62, *p* < .001). Additional correlations are reported in **Table 3** and **Supplemental Figure 1**.

**Table 3.** Convergent Validity: Pearson Correlations (r) between remotely administered Stricker Learning Span measures and in-person administered Auditory Verbal Learning Test measures for all participants. Table used with permission of Mayo Foundation for Medical Education and Research; all rights reserved.

|  | Remotely administered Stricker Learning Span (SLS) |  |  |  |
| --- | --- | --- | --- | --- |
| In-Person AVLT | SLS Sum of Trials | SLS 1-5 Total | SLS Max Span | SLS Delay |
| AVLT Sum of Trials | **0.62** | 0.61 | 0.60 | 0.61 |
| AVLT 1-5 Total | 0.60 | **0.60** | 0.58 | 0.58 |
| AVLT Trial 5 | 0.56 | 0.55 | **0.53** | 0.55 |
| AVLT 30-min Delay | 0.58 | 0.57 | 0.56 | **0.59** |

*Note.* All correlations are significant ( $p$ 's <0.001). Correlations in bold show the relationship between the most similar AVLT and MTD measures. AVLT = Auditory Verbal Learning Test; AVLT Sum of Trials = AVLT 1-5 total + Trial 6 + 30-minute delay; SLS = Stricker Learning Span; SLS Max Span = maximum number of words recognized across any of the 5 learning trials; SLS 1-5 Total = sum of words correctly recognized across trials 1-5; SLS Sum of Trials = SLS 1-5 total + delay. See Supplemental Figure 1 for a full correlation matrix with additional measures. Table used with permission of Mayo Foundation for Medical Education and Research; all rights reserved.

### 3.2 The SLS shows similar ability to differentiate PET-defined biomarker groups compared to the AVLT (Aim 1, all participants)

#### 3.2.1 AUROC comparisons

Total AUROC values for SLS sum of trials vs. AVLT sum of trials were similar for differentiating biomarker groups (*p*’s > .05; **Table 4, Figure 2**). This similarity was seen for all pairwise AUROC comparisons (adjusted and unadjusted models; A- vs A+ and A-T- vs A+T+).

**Table 4.** Logistic regression analysis predicting a) A+ vs. A- and b) A+T+ vs A-T- for both unadjusted models (cognition as the only predictor, on left) and models that also include age, sex, education, in addition to cognition (on right). The *p-*value comparing the AUROCs of the remotely administered Stricker Learning Span and in-person administered Auditory Verbal Learning Test provides a direct comparison of test performance for differentiating biomarker-defined groups.

| **A. Accuracy of SLS and AVL T in Predicting Abnormal Amyloid PET (A+ vs A-)** |  |  |  |  |  |  |  |  |
| --- | --- | --- | --- | --- | --- | --- | --- | --- |
| Characteristic | Unadjusted |  |  |  | Age+Sex+Educ Adjusted |  |  |  |
|  | Odds Ratio<sup>1</sup><br>(95% CI) | *p*-value | AUROC<sup>2</sup><br>(95% CI) | *p*-value<br>comparing<br>AUROCs | Odds Ratio<sup>1</sup><br>(95% CI) | *p*-value | AUROC<sup>2</sup><br>(95% CI) | *p*-value<br>comparing<br>AUROCs |
| **All participants (N=353)** |  |  |  |  |  |  |  |  |
| SLS sum of trials | 0.760 (0.669, 0.860) | <.001 | 0.63 (0.57, 0.70) | 0.76 | 0.966 (0.940, 0.992) | .013 | 0.76 (0.71, 0.81) | 0.90 |
| AVLT sum of trials | 0.741 (0.653, 0.841) | <.001 | 0.64 (0.58, 0.70) |  | 0.965 (0.939, 0.992) | .012 | 0.76 (0.71, 0.81) |  |
| **CU only (N=326)** |  |  |  |  |  |  |  |  |
| SLS sum of trials | 0.753 (0.652, 0.869) | <.001 | 0.63 (0.56, 0.69) | 0.98 | 0.969 (0.940, 1.000) | .013 | 0.77 (0.71, 0.82) | 0.57 |
| AVLT sum of trials | 0.752 (0.653, 0.866) | <.001 | 0.63 (0.57, 0.69) |  | 0.974 (0.945, 1.004) | .012 | 0.76 (0.71, 0.81) |  |
| **B. Accuracy of SLS and AVL T in Predicting Abnormal Amyloid and Tau PET (A+T+ vs A-T-)** |  |  |  |  |  |  |  |  |
| Characteristic | Unadjusted |  |  |  | Age+Sex+Educ Adjusted |  |  |  |
|  | Odds Ratio<sup>1</sup><br>(95% CI) | *p*-value | AUROC<sup>2</sup><br>(95% CI) | *p*-value<br>comparing<br>AUROCs | Odds Ratio<sup>1</sup><br>(95% CI) | *p*-value | AUROC<sup>2</sup><br>(95% CI) | *p*-value<br>comparing<br>AUROCs |
| **All participants (N=353)** |  |  |  |  |  |  |  |  |
| SLS sum of trials | 0.638 (0.536, 0.759) | <.001 | 0.72 (0.64, 0.80) | 0.81 | 0.944 (0.920, 0.970) | <0.001 | 0.84 (0.78, 0.90) | 0.67 |
| AVLT sum of trials | 0.610 (0.503, 0.738) | <.001 | 0.73 (0.65, 0.80) |  | 0.946 (0.921, 0.972) | <0.001 | 0.83 (0.77, 0.90) |  |
| **CU only (N=326)** |  |  |  |  |  |  |  |  |
| SLS sum of trials | 0.659 (0.537, 0.810) | <.001 | 0.69 (0.60, 0.78) | 0.68 | 0.960 (0.931, 0.988) | .007 | 0.83 (0.77, 0.90) | 0.26 |
| AVLT sum of trials | 0.679 (0.545, 0.847) | <.001 | 0.67 (0.59, 0.76) |  | 0.971 (0.942, 1.001) | .059 | 0.81 (0.74, 0.88) |  |

*Note:* The biomarker negative group is the reference group (e.g., A- vs A+ for part A; A-T- vs A+T+ for part B). AUROC = area under the receiver operating characteristic curve; AVL T = Rey's Auditory Verbal Learning Test; AVL T sum of trials = trials 1-5 total correct + trial 6 short-delay correct + 30-minute delay correct, in raw score units. SLS = Stricker Learning Span; SLS sum of trials = 1-5 correct + delay correct, in raw score units. Table used with permission of Mayo Foundation for Medical Education and Research; all rights reserved.
<sup>1</sup> Note that SLS and AVLT odds ratios cannot be compared directly since these two scores are on a different scale (raw scores are used). AUROC data are the focus of test comparisons. Lower test performance is associated with higher odds of being in the biomarker positive group. For example, for the A-T- vs A+T+ comparison (all participants, unadjusted model OR=0.638), each 10-point decrease in SLS sum of trials is associated with 57% increased odds of being A+T+ (95% CI 32% - 87%) calculated as $e^{-1 \cdot \ln(OR)}$ .
<sup>2</sup> Note, for yes/no responses the AUROC is equivalent to concordance, the probability that a randomly selected participant with a yes outcome (positive biomarker group) will have a larger predicted probability than a randomly selected participant with a no outcome (negative biomarker group).

**Figure 2.**
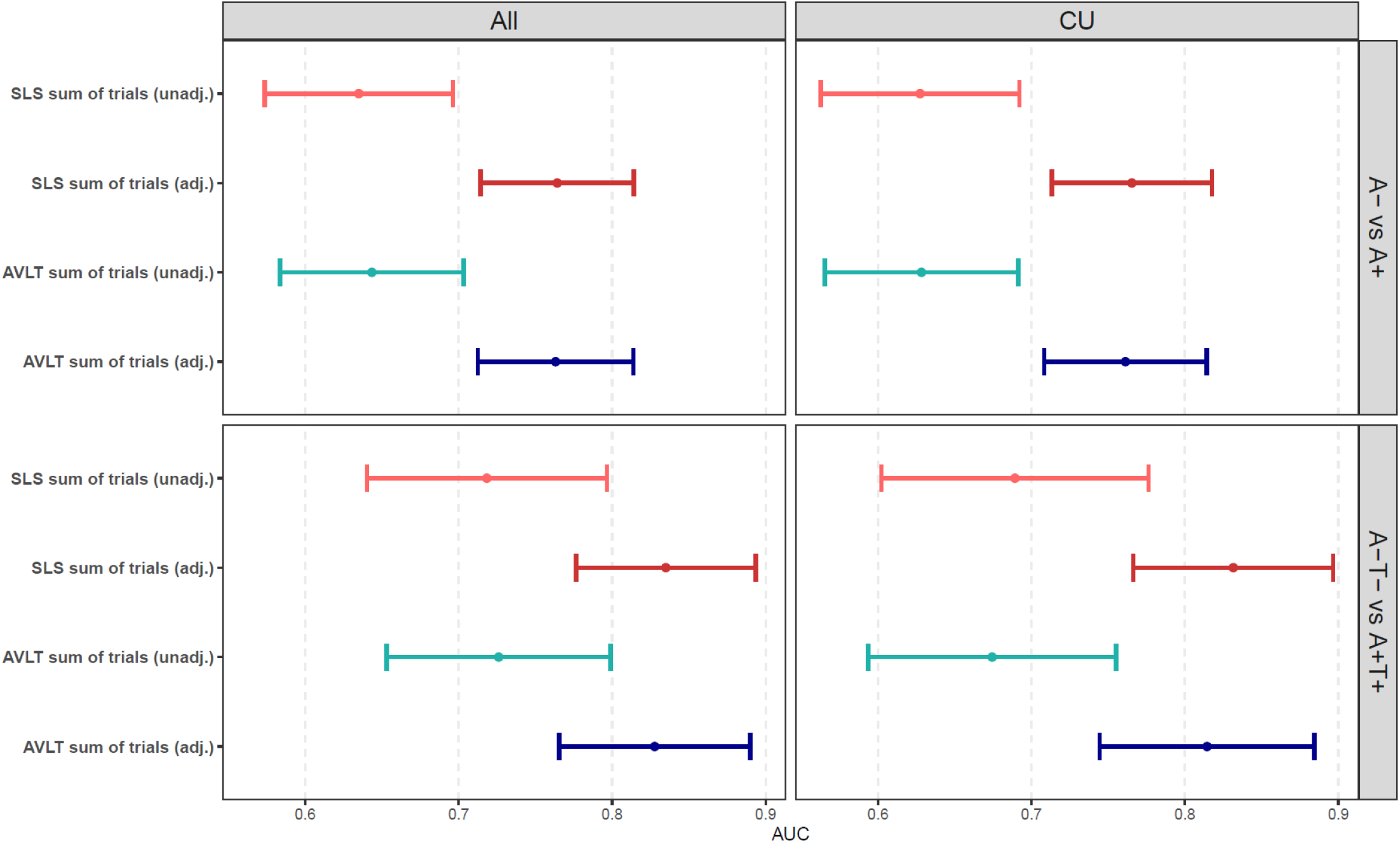
AUROC (95% CI) values for remotely-administered SLS sum of trials and in-person administered AVLT sum of trials. *Note*. All models significantly differentiate biomarker groups better than chance (no AUROC confidence intervals include 0.5). Stricker Learning Span (SLS) sum of trials = 1-5 total correct + delay. Auditory Verbal Learning Test (AVLT) sum of trials = 1-5 total + Trial 6 + 30-minute delay recall. Unadj. = unadjusted models. Adj. = model adjusts for age, education and sex. Figure used with permission of Mayo Foundation for Medical Education and Research; all rights reserved.

##### 3.2.1a Amyloid Groups

Unadjusted models using only the primary cognitive variable as the predictor show that both the SLS and AVLT significantly differentiate A- vs A+ (AUROCs of 0.63 and 0.64, respectively). Adjusted models that include demographic variables increase the overall AUROC values of the full model (both AUROCs=0.76), and both the SLS and AVLT significantly improve biomarker group prediction over and above the demographic variables.

##### 3.2.1b Amyloid and Tau Groups

Unadjusted models using only the primary cognitive variable as the predictor show that both the SLS and AVLT significantly differentiate individuals without AD biomarkers (A-T-) from those with biological AD (A+T+; AUROCs of 0.72-0.73). Adjusted models that include demographic variables increase the overall AUROC values of the full model (AUROCs of 0.83-0.84), and both the SLS and AVLT significantly improve biomarker group prediction over and above the demographic variables.

#### 3.2.2 Descriptive effect sizes from group difference analyses (**Table 5**)

##### 3.2.2a Amyloid Groups

Unadjusted analyses showed medium group effect sizes for both SLS sum of trials (g = -0.51) and AVLT sum of trials (g = -0.55) for differentiating A- and A+ groups (**Figure 3**). Effect sizes after adjusting for demographics were decreased and small in magnitude (g’s = -0.26 to -0.27), and group differences remained significant.

**Table 5.** Unadjusted and adjusted Hedge’s g effect sizes (95% CI) between biomarker subgroup for all variables.

|  | A- vs A+ |  |  |  | A-T- vs A+T+ |  |  |  |
| --- | --- | --- | --- | --- | --- | --- | --- | --- |
|  | All A participants (N=353) |  | CU A participants (N=326) |  | All AT participants (N=259) |  | CU AT participants (N=224) |  |
|  | Unadj g | Adj g | Unadj g | Adj g | Unadj g | Adj g | Unadj g | Adj g |
| **Primary variables** |  |  |  |  |  |  |  |  |
| SLS sum of trials | -0.51*<br>(-0.73, -0.28) | -0.26*<br>(-0.47, -0.05) | -0.47*<br>(-0.71, -0.24) | -0.22<br>(-0.44, 0.00) | -0.88*<br>(-1.19, -0.57) | -0.53*<br>(-0.78, -0.28) | -0.74*<br>(-1.08, -0.39) | -0.37*<br>(-0.63, -0.10) |
| AVLT sum of trials | -0.55*<br>(-0.77, -0.33) | -0.27*<br>(-0.48, -0.06) | -0.48*<br>(-0.72, -0.25) | -0.18<br>(-0.40, 0.04) | -0.88*<br>(-1.19, -0.57) | -0.51*<br>(-0.76, -0.25) | -0.62*<br>(-0.96, -0.28) | -0.25<br>(-0.52, 0.01) |
| **Secondary variables** |  |  |  |  |  |  |  |  |
| SLS Max Span | -0.45*<br>(-0.67, -0.23) | -0.21<br>(-0.42, 0.00) | -0.40*<br>(-0.64, -0.17) | -0.16<br>(-0.37, 0.06) | -0.81*<br>(-1.12, -0.50) | -0.47*<br>(-0.73, -0.22) | -0.65*<br>(-0.99, -0.30) | -0.30*<br>(-0.56, -0.03) |
| AVLT Trial 5 | -0.46*<br>(-0.68, -0.24) | -0.18<br>(-0.39, 0.03) | -0.41*<br>(-0.64, -0.17) | -0.12<br>(-0.34, 0.10) | -0.74*<br>(-1.05, -0.44) | -0.40*<br>(-0.65, -0.15) | -0.53*<br>(-0.87, -0.19) | -0.19<br>(-0.46, 0.07) |
| SLS 1-5 Total | -0.50*<br>(-0.72, -0.28) | -0.26*<br>(-0.47, -0.05) | -0.47*<br>(-0.71, -0.24) | -0.23*<br>(-0.44, -0.01) | -0.86*<br>(-1.16, -0.55) | -0.51*<br>(-0.76, -0.26) | -0.73*<br>(-1.07, -0.38) | -0.36*<br>(-0.63, -0.10) |
| AVLT 1-5 Total | -0.54*<br>(-0.76, -0.32) | -0.28*<br>(-0.49, -0.07) | -0.48*<br>(-0.72, -0.25) | -0.21<br>(-0.43, 0.00) | -0.82*<br>(-1.13, -0.51) | -0.48*<br>(-0.73, -0.23) | -0.59*<br>(-0.93, -0.25) | -0.25<br>(-0.52, 0.01) |
| SLS Delay | -0.48*<br>(-0.70, -0.26) | -0.25*<br>(-0.46, -0.04) | -0.42*<br>(-0.65, -0.19) | -0.18<br>(-0.39, 0.04) | -0.88*<br>(-1.19, -0.57) | -0.54*<br>(-0.79, -0.29) | -0.69*<br>(-1.03, -0.35) | -0.34*<br>(-0.60, -0.07) |
| AVLT 30-min delay | -0.48*<br>(-0.71, -0.26) | -0.20<br>(-0.41, 0.01) | -0.40*<br>(-0.63, -0.17) | -0.10<br>(-0.32, 0.12) | -0.86*<br>(-1.17, -0.55) | -0.50*<br>(-0.75, -0.25) | -0.56*<br>(-0.90, -0.22) | -0.22<br>(-0.48, 0.04) |
| **Exploratory variables** |  |  |  |  |  |  |  |  |
| SLS Retention | -0.31*<br>(-0.53, -0.09) | -0.24*<br>(-0.45, -0.03) | -0.22<br>(-0.45, 0.01) | -0.14<br>(-0.36, 0.08) | -0.55*<br>(-0.85, -0.24) | -0.40*<br>(-0.65, -0.15) | -0.36*<br>(-0.70, -0.03) | -0.22<br>(-0.48, 0.05) |
| AVLT Retention | -0.41*<br>(-0.63, -0.19) | -0.21<br>(-0.42, 0.00) | -0.27*<br>(-0.50, -0.04) | -0.08<br>(-0.30, 0.14) | -0.79*<br>(-1.09, -0.48) | -0.50*<br>(-0.75, -0.25) | -0.36*<br>(-0.70, -0.02) | -0.15<br>(-0.41, 0.11) |
| AVLT Trial 6 | -0.51*<br>(-0.73, -0.28) | -0.20<br>(-0.41, 0.01) | -0.42*<br>(-0.66, -0.19) | -0.11<br>(-0.33, 0.11) | -0.85*<br>(-1.16, -0.54) | -0.46*<br>(-0.71, -0.21) | -0.59*<br>(-0.93, -0.25) | -0.22<br>(-0.48, 0.04) |
| AVLT Recogn PC | -0.47*<br>(-0.69, -0.25) | -0.28*<br>(-0.50, -0.07) | -0.37*<br>(-0.60, -0.14) | -0.17<br>(-0.39, 0.04) | -0.77*<br>(-1.08, -0.46) | -0.49*<br>(-0.74, -0.24) | -0.43*<br>(-0.77, -0.09) | -0.17<br>(-0.44, 0.09) |

\* $p < .05$
*Note:* The most similar AVLT and MTD measures and grouped together to facilitate qualitative comparison; note that the retention variables should not be directly compared due to fundamental differences in test design that is expected to yield discrepant results (e.g., randomization of SLS word list presentation to minimize recency effect on leaning trials). Table used with permission of Mayo Foundation for Medical Education and Research; all rights reserved.

**Figure 3.**
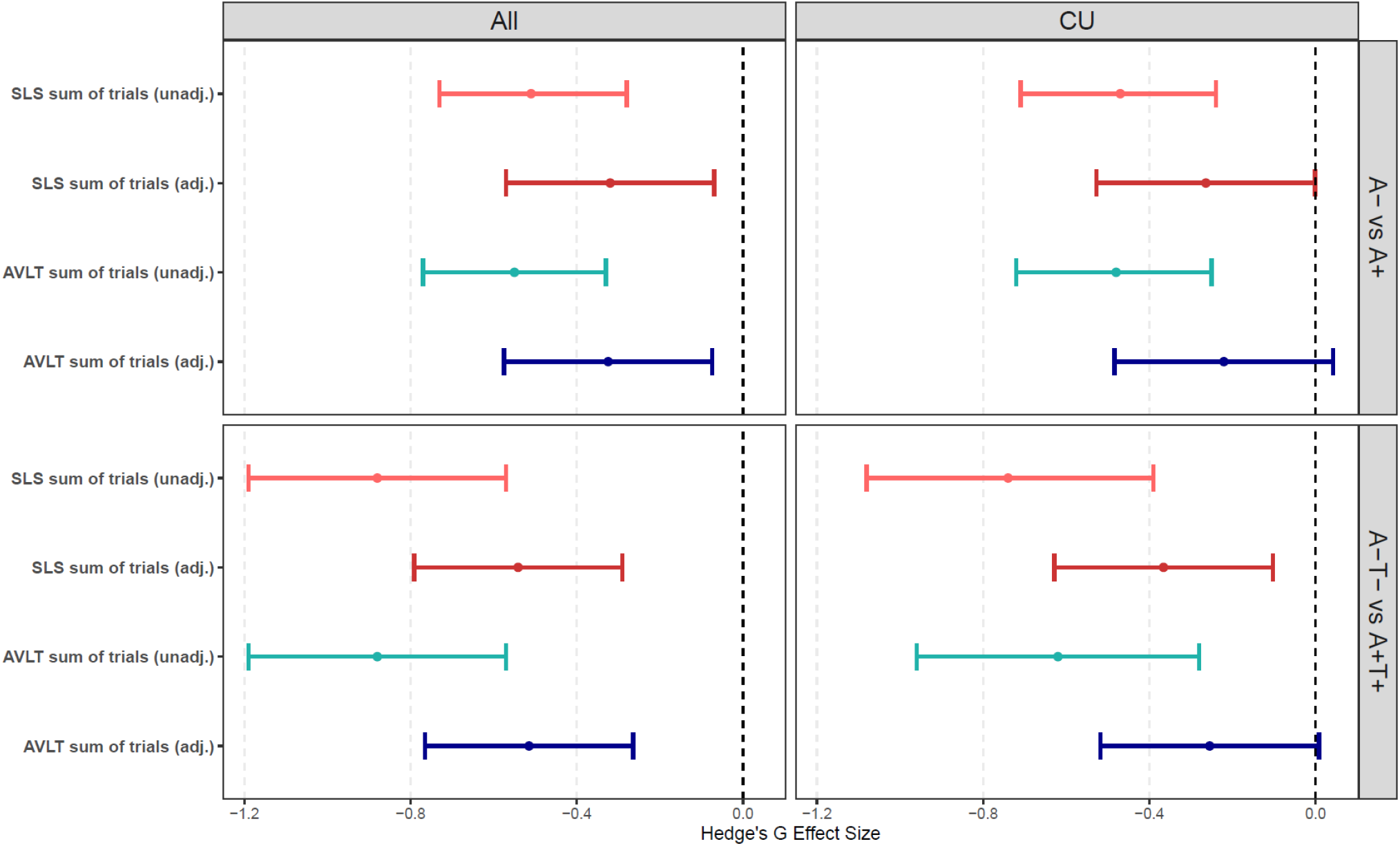
Hedge’s g effect sizes with confidence intervals to show the magnitude of group differences for remotely administered Stricker Learning Span (SLS, red shades) and in-person Auditory Verbal Learning Test (AVLT) measures (blue shades) across biomarker groups (A- vs A+ top; A-T- vs A+T+ bottom) in all participants (left) and Cognitively Unimpaired participants (right). *Note*. Groups do not significantly differ when the CI includes 0 (dashed line). Unadj. = unadjusted models. Adj. = model adjusts for age, education and sex. Remote SLS Sum of Trials = SLS trials 1-5 correct + delay correct. In-person AVLT Sum of Trials = AVLT 1-5 correct + Trial 6 (short-delay) correct + 30-minute recall correct). See Tables 4 and 5 for direction of effect sizes and confidence intervals. Figure used with permission of Mayo Foundation for Medical Education and Research; all rights reserved.

##### 3.2.2b Amyloid and Tau Groups

Unadjusted analyses showed large group difference effect sizes for SLS sum of trials and AVLT sum of trials (both g’s = -0.88) for differentiating A- T- and A+T+ groups (**Figure 3**). Effect sizes after adjusting for demographics were decreased but remained moderate in magnitude and significant for both SLS sum of trials (g = -0.53) and AVLT sum of trials (g = -0.51).

### 3.3 The SLS shows similar ability to differentiate PET-defined biomarker groups compared to the AVLT for preclinical AD (CU participants only; Aim 2)

Because the majority (93%) of participants in the overall sample are CU, limiting analyses only to CU participants had relatively little impact on the results. Findings show that the SLS is sensitive to preclinical AD, consistent with our Aim 2 hypothesis.

#### 3.3.1 AUROC comparisons

Total AUROC values for SLS sum of trials vs. AVLT sum of trials were similar for differentiating biomarker groups in CU participants (*p*’s > .05 for each pairwise comparison; **Table 4**).

##### 3.3.1a Amyloid Groups

Unadjusted models using only the primary cognitive variable as the predictor show that both the SLS and AVLT significantly differentiate A- vs A+ (both AUROCs = 0.63). Adjusted models that include demographic variables increase the overall AUROC values of the full model (AUROCs=0.76-0.77), and both the SLS and AVLT significantly improve biomarker group prediction over and above the demographic variables.

##### 3.3.1b Amyloid and Tau Groups

Unadjusted models using only the primary cognitive variable as the predictor show that both the SLS and AVLT significantly differentiate individuals without AD biomarkers (A-T-) from those with biological AD (A+T+; AUROCs = 0.67-0.69).

Adjusted models that include demographic variables increase the overall AUROC values of the full model (AUROCs = 0.81-0.83). The SLS significantly improved biomarker group prediction over and above the demographic variables; the AVLT approached significance (*p*=0.06).

#### 3.3.2 Descriptive effect sizes from group difference analyses (**Table 5**)

##### 3.3.2a Amyloid Groups

Unadjusted analyses showed small to medium group effect sizes for both SLS sum of trials (g = -0.47) and AVLT sum of trials (g = -0.48) for differentiating A- and A+ groups (**Figure 3**). Effect sizes after adjusting for demographics were decreased and small in magnitude (g’s = -0.18 to -0.22), and group differences did not reach significance.

##### 3.3.2b Amyloid and Tau Groups

Unadjusted analyses showed medium group difference effect sizes for SLS sum of trials (g = -0.74) and AVLT sum of trials (g = -0.62) for differentiating A-T- and A+T+ groups (**Figure 3**). Effect sizes after adjusting for demographics were decreased (small in magnitude) with SLS sum of trials remaining significant (g = -0.37) and AVLT sum of trials no longer reaching significance (g = -0.25).

### 3.4 Learning measures show similar ability as delay memory measures to differentiate A-T- and A+T+ groups for both the SLS and the AVLT

#### 3.4.1 All participants

All secondary SLS and AVLT variables show results in the expected direction with lower performance in the A+T+ compared to A-T-group (p’s < .05 for both unadjusted and adjusted analyses), with generally similar effect sizes across comparable SLS and AVLT variable pairs (**Table 5**). Within-test descriptive comparisons show that learning variables (1-5 total) show effect sizes that are similar in magnitude as delay variables. For example, SLS 1-5 (g = -.86) and delay (g = -.88) both show large unadjusted effect sizes, and AVLT 1-5 (g = -.82) and delay (g = -.86) also show large unadjusted effect sizes (**Figure 4**). Trials 1-5 total (g=-.86 SLS and -.82 AVLT) may be a slightly more advantageous learning measure for group discrimination relative to SLS max span (−.81) or AVLT Trial 5 (−.74). Similarly, comparison of two different types of delayed memory measures suggests a slight advantage for delay total correct relative to retention (−.88 vs -.55 for SLS, respectively and -.86 vs -.79 for AVLT, respectively). See **Figure 5** for a visualization of trial-by-trial data for both the SLS and AVLT. See **Supplemental Tables 1 and 2** for results of other memory tests.

**Figure 4.**
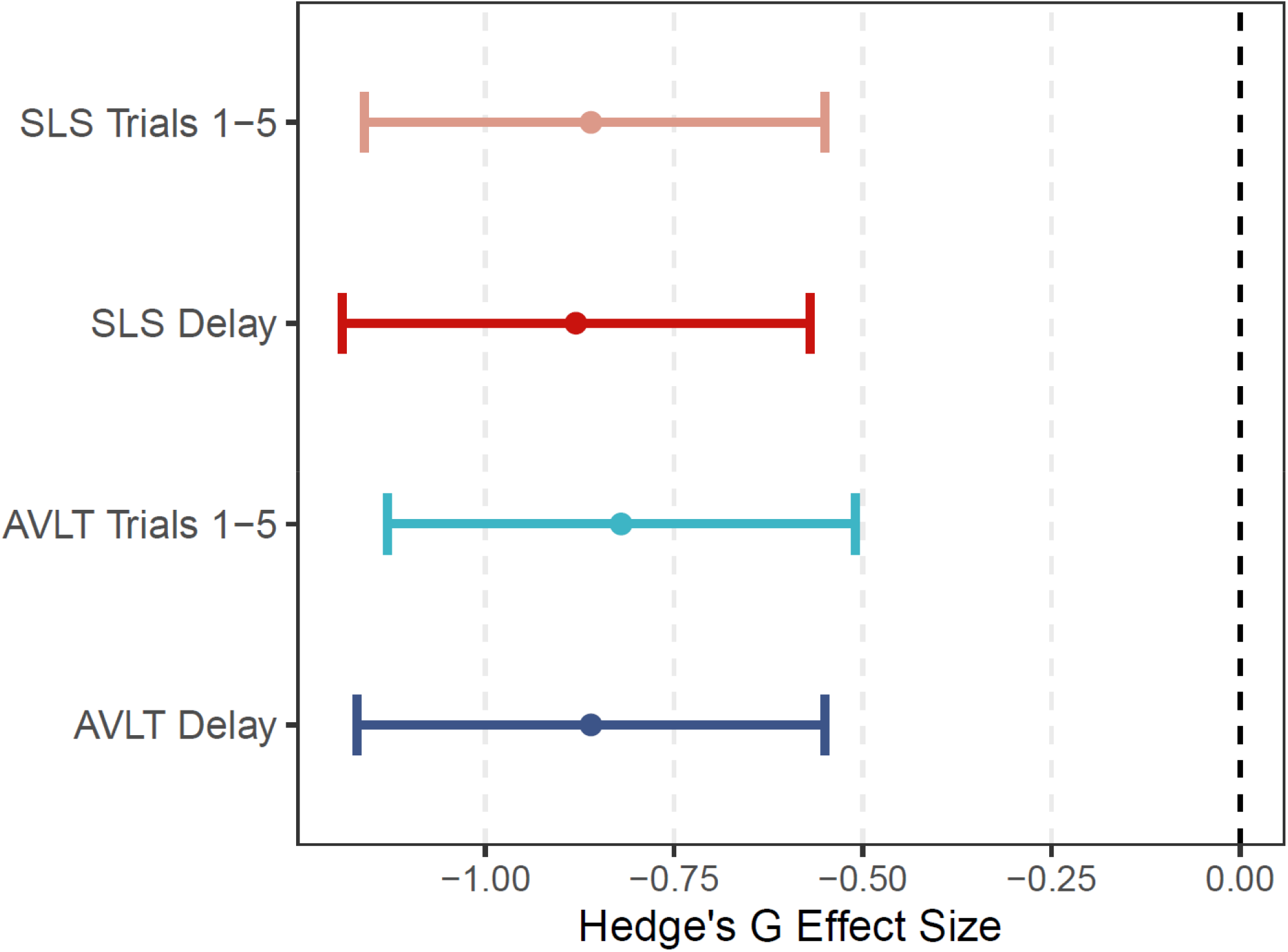
Hedge’s g unadjusted effect sizes comparing ability of learning and delay trials to differentiate individuals without AD biomarkers from those with biological AD (A-T- vs A+T+). *Note*. Stricker Learning Span (SLS) Trials 1-5 = trials 1-5 total correct; Auditory Verbal Learning Test (AVLT) Trials 1-5 = 1-5 total correct; AVLT Delay = 30-minute delayed recall. Figure used with permission of Mayo Foundation for Medical Education and Research; all rights reserved.

**Figure 5.**
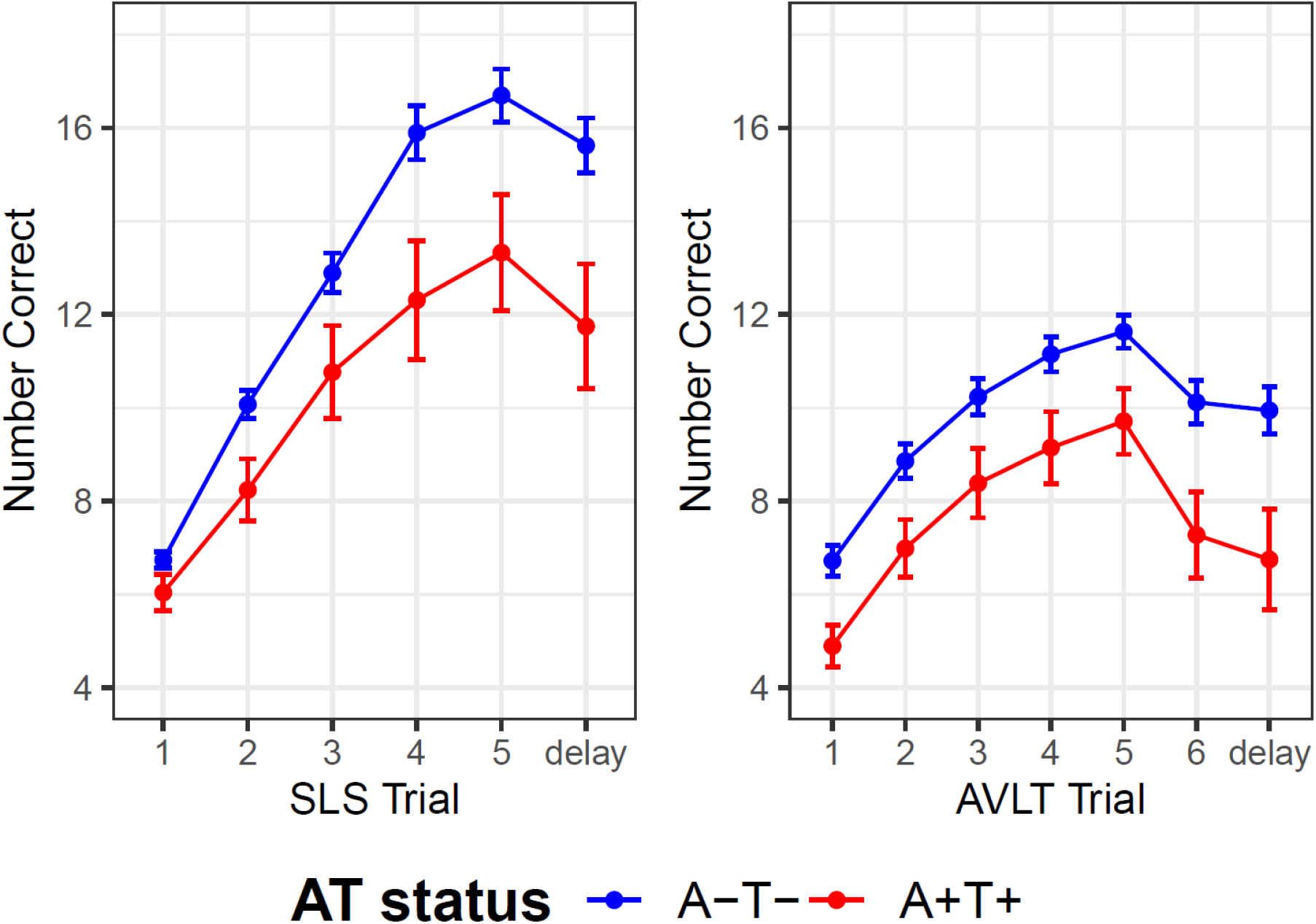
Stricker Learning Span (SLS, left panel) and Auditory Verbal Learning Test (AVLT, right panel) learning slopes and delayed memory performances across all participants without AD biomarkers (A-T-, blue) and all participants with biological AD (A+T+, red). *Note*. Sum of trials is the primary variable for each test; these figures show the data comprising sum of trials for each measure (sum of trials = total correct items for all trials displayed in each respective figure). SLS items presented differ by trial based on computer adaptive testing rules, thus the highest possible number of presented across trials vary (see Figure 1 for ceiling and floor values for Trials 1-5; the delay trial can range from 8-23). The SLS uses 4-choice recognition to test item memory. In contrast, 15 words are presented each time for AVLT trials (constant range of 0-15) and item memory is tested with free recall. See Supplemental Table 2 for numeric results. AVLT Trial 6 = short-delay. AVLT delay = 30-minute delay. Figure used with permission of Mayo Foundation for Medical Education and Research; all rights reserved.

##### Data Availability

The data that support the findings of this study are available from the corresponding author upon reasonable request and with appropriate institutional approvals.

## 4. Discussion

This study followed a novel approach to test validation and established the criterion validity of an unsupervised computer adaptive word list memory test (SLS) completed outside of a clinic setting. The SLS differentiates AD biomarker-defined groups as well as a traditional word list recall test (AVLT) administered by trained psychometrists in a clinic setting. Specifically, our Aim 1 hypothesis was supported by AUROC comparisons that showed remotely administered SLS sum of trials and in-person-administered AVLT sum of trials have comparable ability to differentiate individuals on the Alzheimer’s continuum (A+) or not (A-) and individuals meeting a research framework for a biological diagnosis of AD (A+T+) or not (A-T-) in a predominantly cognitively unimpaired sample.

In line with our prior results showing that the AVLT has the potential to be useful for detecting subtle objective cognitive decline in preclinical AD ^33^, our current results extend this prior AVLT finding and suggest that the SLS also has promise in this regard (Aim 2). Specifically, when limiting the sample to CU participants, our AUROC results show that the SLS by itself could help predict, better than chance, which individuals had elevated brain amyloid vs. did not and which had elevated brain amyloid and tau vs did not. In contrast, our prior work examining the utility of the Learning/Working Memory index comprised of visual recognition and working memory tasks from the Cogstate Brief Battery administered in clinic did not significantly differentiate biomarker groups better than chance (CU A-T- vs CU A+T+ or CU A- T- vs CU A+T-) and showed that the AVLT was significantly better than the Learning/Working Memory Index for differentiating CU A-T- vs CU A+T- when comparing total AUROCs. While the predictive ability of the SLS by itself is relatively modest, predictive ability improves when demographic variables are added to the model and the SLS continues to show an independent effect over and above demographic variables. For example, a model with age, sex, education and SLS sum of trials together had an AUROC of 0.83 for predicting A+T+ status in CU participants. Thus, our current results suggest that the SLS could be a scalable, easily accessible addition to a multivariable model approach that improves overall prediction of AD risk. Given its capacity for remote self-administration, the SLS would be a good candidate screening measure in non-specialty care settings to use in combination with plasma biomarkers to inform the need for further work-up.

The highly overlapping results for the ability of the SLS and AVLT to discriminate AD biomarker groups are particularly interesting given that although the SLS was designed to mimic the sensitivity of the AVLT, it was not designed to be a one-to-one adaptation of the AVLT. Results of correlation analyses align with this intent and support our hypothesis that the SLS and AVLT would show a significant correlation (r = 0.62 observed), and therefore further support convergent validity (Aim 3). Our initial pilot study similarly supported the convergent validity of the SLS, but slightly lower correlation coefficients were observed in that study, likely due to homogeneity of that sample (e.g., all female, restricted age range, excluded individuals with dementia), and potentially due to a longer duration between in-person and remote testing (average 10 months)^13^. The current results are particularly notable given that an exact computerized replication of the AVLT facilitated by audio recording and speech recognition to allow self-administered on an iPad showed only slightly higher correlations (r=0.63-0.70) with the AVLT as typically administered in a well-controlled cross-over design completed in a clinic setting^43^.

We also qualitatively compared commonly derived supraspan wordlist indices within each test given that the learning indices may have equivalent utility for the early detection of AD as delayed memory indices^44,45^. Results supported our hypothesis that word list learning measures would show similar sensitivity as word list delay memory measures to biologically defined AD (A-T- vs A+T+; Aim 4). The effect sizes of learning trials and delayed memory are similar (**Figure 4**). We chose to focus on delay items correct instead of percent retention (i.e., savings) for the comparison to learning indices. It is important to note that retention is largely dependent on specific test design characteristics including the influence of serial position effects^46-48^. The SLS randomizes word order to minimize the recency effect often observed in individuals with Alzheimer’s dementia that leads to an over-estimate of true forgetting^49^. For example, learning to criterion studies demonstrate that individuals in the early stages of AD take a longer time to reach criterion, reflective of lower learning ability, whereas once learning is equated by reaching criterion rates of forgetting are similar to healthy control participants^48,50,51^. Accordingly, SLS learning indices (1-5, max span) demonstrate a larger effect size than SLS retention in biologically defined AD versus those without AD biomarkers, whereas AVLT retention effect size is more similar to AVLT learning indices.

This study has several strengths. Most studies examining the ability of cognitive measures to differentiate biomarker groups have focused on differentiation of A+ versus A-individuals^52,53^. Our inclusion of tau status defined by PET imaging, in addition to amyloid, is a strength. Our population-based sample helps to increase generalizability to clinical settings where comorbidities are common. Our approach of reporting both unadjusted and adjusted effect sizes helps illustrate the robust biomarker-group difference effect sizes observed in unadjusted analyses. For example, the SLS showed a large group difference effect size across A-T- and A+T+ groups in all participants (−0.88), that decreased to a medium effect size when controlling for demographics (−0.53) and was further attenuated when limited to CU only (−0.74 unadjusted, -0.37 adjusted). There is growing evidence that cognitive decline is not a normal part of the aging process, but rather, is reflective of previously undetected neuropathologies that increase in prevalence with increasing age^54-56^. To maximize the utility of cognitive measures for informing risk of AD biomarker positivity, we recommend the use of raw scores and argue that the effect of age should not be routinely “adjusted” away as it decreases the predictive power of cognition.

Several limitations should also be noted. First, because this is a population-based study, the racial and ethnic characteristics reflect that of Olmsted County from which participants are randomly sampled, resulting in a predominantly White, Non-Hispanic sample. Second, the predominantly CU composition of this sample may have decreased AUROC values and magnitude of effect sizes for differentiating biomarker positive and negative groups, as suggested by the highly similar results seen when limiting analyses only to CU participants. A more balanced sample design, with more inclusion of individuals with mild to moderate dementia, could suggest greater utility of these memory measures for identifying individuals at risk for biomarker positivity; the current results are more relevant for preclinical detection given that 93% of our sample is CU. Third, a majority of the sample (83%) had prior exposure to the AVLT given the longitudinal nature of the MCSA and ADRC studies, thus practice effects could have impacted the ability of the AVLT to discriminate biomarker groups. Because biomarker negative participants benefit more from practice effects than biomarker positive participants, it is possible this could have amplified group difference effects for the AVLT^15,57^. Future work is needed to replicate these results in a setting where both the SLS and AVLT are baseline administrations. Similarly, the entirely unsupervised and remote approach for the SLS could dampen the sensitivity of the SLS as the results presented in this study include all available remote data. While we capture participant-reported information about test interference, noise in the test environment, and participant comments that can provide additional information about test interruptions or environmental considerations, because our goal was to establish the robust criterion validity of the SLS “in the wild,” we did not apply any exclusionary criterion based on this information in the present study. Another reason we did not want to prematurely apply such exclusionary criteria is that individuals who are less able to follow instructions provided to establish the recommended test environment may be more likely to have cognitive impairment. Thus, increased likelihood of lower test performance in an uncontrolled environment, worsened by environmental distractions, could also be related to risk of cognitive decline. If cognitive screening / risk for cognitive decline is the goal, worse performance in remote settings may help identify risk in a way not captured by controlled clinical settings, adding an element of ecological validity in terms of ability to adapt to a new task without assistance. Future work will examine whether and to what degree these factors may influence test performance, as increased distractions in the home environment can negatively impact performance^58^. Finally, while use of a strictly biomarker-defined ground truth is a novel aspect of this study, in vivo PET biomarkers also have some limitations such as high but imperfect reliability (manifested by “noise” in the trajectories of imaging results over time in some individuals), and the fact that PET measures of amyloid and tau pathology have a sensitivity floor and medically significant pathology can exist that lies beneath this detection threshold^59^. Also, we adopted a liberal window for inclusion of available biomarker data to allow for some missed scanning opportunities during the COVID-19 pandemic, to maximize the sample size and because of the generally good stability of amyloid and tau classifications^60^, but this also decreases study precision relative to a narrower time window.

In summary, SLS test design prioritized remote assessment needs and a computer-adaptive approach^12^. Even though the SLS is not a direct adaptation of the AVLT, our results show highly similar ability of the remotely-administered SLS and in-person-administered AVLT to differentiate AD biomarker-defined groups. These results challenge preconceived notions about memory assessment by showing that creative use of a recognition memory paradigm that emphasizes learning in an all-remote unsupervised sample differentiates AD biomarker-defined groups as effectively as a traditional word list memory measure based on free recall responses.

## Supporting information

Supplemental Online Resources

## Acknowledgment

The authors wish to thank the participants and staff at the Mayo Clinic Study of Aging and Mayo Alzheimer’s Disease Research Center. Research reported in this publication was supported by the Kevin Merszei Career Development Award in Neurodegenerative Diseases Research IHO Janet Vittone, MD, the Rochester Epidemiology Project (R01 AG034676), the National Institute on Aging of the National Institutes of Health (grant numbers R21AG073967, P30 AG062677, P50 AG016574, U01 AG006786, RF1 AG55151, R01 AG041851, R37 AG011378), the Robert Wood Johnson Foundation, The Elsie and Marvin Dekelboum Family Foundation, GHR Foundation, Alzheimer’s Association, and the Mayo Foundation for Education and Research. The content is solely the responsibility of the authors and does not necessarily represent the official views of the National Institutes of Health. A Mayo Clinic invention disclosure has been submitted for the Stricker Learning Span and the Mayo Test Drive platform (NHS, JLS). We have no other conflicts of interest to disclose related to this work.

The following disclosures are present, outside the scope of this work: NHS has received research support from NIH and Biogen. MMMi has consulted for Biogen, Eli Lilly, Lab Corp, Merck, and Siemens Healthineers and receives research support from NIH. MMMa and JAF receive research support from NIH. DSK serves on a Data Safety Monitoring Board for the DIAN-TU study and is an investigator in clinical trials sponsored by Lilly Pharmaceuticals, Biogen, and the University of Southern California. RCP has served as a consultant for Hoffman-La Roche Inc., Merck Inc., Genentech Inc., Biogen Inc., Eisai, Inc. and Nestle, Inc. He also receives NIH support. CRJ serves on an independent data monitoring board for Roche but he receives no personal compensation from any commercial entity. CRJ receives research support from NIH and the Alexander Family Alzheimer’s Disease Research Professorship of the Mayo Clinic.

## References

1 Cummings, J., Lee, G., Zhong, K., Fonseca, J. & Taghva, K. Alzheimer’s disease drug development pipeline: 2021. Alzheimers Dement (N Y) 7, e12179, doi:10.1002/trc2.12179 (2021).

2 Dorsey, E. R., Kluger, B. & Lipset, C. H. The New Normal in Clinical Trials: Decentralized Studies. Ann. Neurol. 88, 863–866, doi:10.1002/ana.25892 (2020).

3 Sabbagh, M. et al. Early Detection of Mild Cognitive Impairment MCI in an At Home Setting. J Prev Alzheimers Dis, doi:10.14283/jpad.2020.21 (2020).

4 Sabbagh, M. N. et al. Rationale for Early Diagnosis of Mild Cognitive Impairment (MCI) Supported by Emerging Digital Technologies. J Prev Alzheimers Dis 7, 158–164, doi:10.14283/jpad.2020.19 (2020).

5 Bauer, R. M. & Bilder, R. M. in APA Handbook of Neuropsychology Vol. 2: Neuroscience and Neuromethods (eds Gregory G. Brown, Tricia Z. King, Kathleen Y. Haaland, & Bruce Crosson) (American Psychological Association, 2023, in press).

6 Mackin, R. S. et al. Reliability and Validity of a Home-Based Self-Administered Computerized Test of Learning and Memory Using Speech Recognition. Neuropsychol. Dev. Cogn. B Aging Neuropsychol. Cogn., 1–15, doi:10.1080/13825585.2021.1927961 (2021).

7 Marra, D. E., Hamlet, K. M., Bauer, R. M. & Bowers, D. Validity of teleneuropsychology for older adults in response to COVID-19: A systematic and critical review. Clin. Neuropsychol., 1–42, doi:10.1080/13854046.2020.1769192 (2020).

8 Cromer, J. A. et al. Comparison of Cognitive Performance on the Cogstate Brief Battery When Taken In-Clinic, In-Group, and Unsupervised. Clin. Neuropsychol. 29, 542–558, doi:10.1080/13854046.2015.1054437 (2015).

9 Mielke, M. M. et al. Performance of the CogState computerized battery in the Mayo Clinic Study on Aging. Alzheimer’s & Dementia 11, 1367–1376, doi:10.1016/j.jalz.2015.01.008 (2015).

10 Stricker, N. H. et al. Longitudinal Comparison of in Clinic and at Home Administration of the Cogstate Brief Battery and Demonstrated Practice Effects in the Mayo Clinic Study of Aging. The journal of prevention of Alzheimer’s disease 7, 21–28, doi:10.14283/jpad.2019.35 (2020).

11 Caselli, R. J. et al. Neuropsychological decline up to 20 years before incident mild cognitive impairment. Alzheimers Dement 16, 512–523, doi:10.1016/j.jalz.2019.09.085 (2020).

12 Stricker, J. L. et al. Neural network process simulations support a distributed memory system and aid design of a novel computer adaptive digital memory test for preclinical and prodromal Alzheimer’s disease. Neuropsychology, doi:10.1037/neu0000847 (2022).

13 Stricker, N. H. et al. A novel computer adaptive word list memory test optimized for remote assessment: Psychometric properties and associations with neurodegenerative biomarkers in older women without dementia. Alzheimers Dement (Amst) 14, e12299, doi:10.1002/dad2.12299 (2022).

14 Lim, Y. Y. et al. Association of deficits in short-term learning and Aβ and hippoampal volume in cognitively normal adults. Neurology, 10.1212/WNL.0000000000010728, doi:10.1212/wnl.0000000000010728 (2020).

15 Machulda, M. M. et al. Practice effects and longitudinal cognitive change in clinically normal older adults differ by Alzheimer imaging biomarker status. Clin. Neuropsychol. 31, 99–117, doi:10.1080/13854046.2016.1241303 (2017).

16 Duff, K. et al. Short-Term Practice Effects and Amyloid Deposition: Providing Information Above and Beyond Baseline Cognition. J Prev Alzheimers Dis 4, 87–92, doi:10.14283/jpad.2017.9 (2017).

17 Bilder, R. M. & Reise, S. P. Neuropsychological tests of the future: How do we get there from here? Clin. Neuropsychol. 33, 220–245, doi:10.1080/13854046.2018.1521993 (2019).

18 Wolters, E. E. et al. Clinical validity of increased cortical uptake of [(18)F]flortaucipir on PET as a biomarker for Alzheimer’s disease in the context of a structured 5-phase biomarker development framework. Eur. J. Nucl. Med. Mol. Imaging 48, 2097–2109, doi:10.1007/s00259-020-05118-w (2021).

19 Chiotis, K. et al. Clinical validity of increased cortical uptake of amyloid ligands on PET as a biomarker for Alzheimer’s disease in the context of a structured 5-phase development framework. Neurobiol. Aging 52, 214–227, doi:10.1016/j.neurobiolaging.2016.07.012 (2017).

20 Jack, C. R., Jr. et al. NIA-AA Research Framework: Toward a biological definition of Alzheimer’s disease. Alzheimers Dement 14, 535–562, doi:10.1016/j.jalz.2018.02.018 (2018).

21 St Sauver, J. L. et al. Data resource profile: the Rochester Epidemiology Project (REP) medical records-linkage system. Int. J. Epidemiol. 41, 1614–1624, doi:10.1093/ije/dys195 (2012).

22 Roberts, R. O. et al. The Mayo Clinic Study of Aging: Design and sampling, participation, baseline measures and sample characteristics. Neuroepidemiology 30, 58–69, doi:10.1159/000115751 (2008).

23 Kokmen, E., Smith, G. E., Petersen, R. C., Tangalos, E. & Ivnik, R. C. The short test of mental status: Correlations with standardized psychometric testing. Arch. Neurol. 48, 725–728, doi:10.1001/archneur.1991.00530190071018 (1991).

24 Morris, J. C. The Clinical Dementia Rating (CDR): Current version and scoring rules. Neurology 43, 2412–2414, doi:10.1212/WNL.43.11.2412-a (1993).

25 Petersen, R. C. Mild cognitive impairment as a diagnostic entity. J. Intern. Med. 256, 183–194, doi:10.1111/j.1365-2796.2004.01388.x. (2004).

26 American Psychiatric Association. Diagnostic and Statistical Manual of Mental Disorders (DSM-IV). 4th edn, (American Psychiatric Association, 1994).

27 Jack, C. R., Jr. et al. 11C PiB and structural MRI provide complementary information in imaging of Alzheimer’s disease and amnestic mild cognitive impairment. Brain 131, 665–680 (2008).

28 Jack, C. R., Jr. et al. Defining imaging biomarker cut points for brain aging and Alzheimer’s disease. Alzheimers Dement 13, 205–216, doi:10.1016/j.jalz.2016.08.005 (2017).

29 Vemuri, P. et al. Tau-PET uptake: Regional variation in average SUVR and impact of amyloid deposition. Alzheimers Dement (Amst) 6, 21–30, doi:10.1016/j.dadm.2016.12.010 (2017).

30 Klunk, W. E. et al. The Centiloid Project: standardizing quantitative amyloid plaque estimation by PET. Alzheimers Dement 11, 1-15 e11-14, doi:10.1016/j.jalz.2014.07.003 (2015).

31 Alden, E. C. et al. Diagnostic accuracy of the Cogstate Brief Battery for prevalent MCI and prodromal AD (MCI A(+) T(+)) in a population-based sample. Alzheimers Dement 17, 584–594, doi:10.1002/alz.12219 (2021).

32 Pudumjee, S. B. et al. A Comparison of Cross-Sectional and Longitudinal Methods of Defining Objective Subtle Cognitive Decline in Preclinical Alzheimer’s Disease Based on Cogstate One Card Learning Accuracy Performance. J. Alzheimers Dis. 83, 861–877, doi:10.3233/JAD-210251 (2021).

33 Stricker, N. H. et al. Diagnostic and Prognostic Accuracy of the Cogstate Brief Battery and Auditory Verbal Learning Test in Preclinical Alzheimer’s Disease and Incident Mild Cognitive Impairment: Implications for Defining Subtle Objective Cognitive Impairment J. Alzheimers Dis. 76, 261–274, doi:10.3233/JAD-200087 (2020).

34 Stricker, N. H. et al. Mayo Normative Studies: Regression-Based Normative Data for the Auditory Verbal Learning Test for Ages 30–91 Years and the Importance of Adjusting for Sex. J. Int. Neuropsychol. Soc., 1–16, doi:10.1017/S1355617720000752 (2020).

35 Ferman, T. J. et al. Mayo’s Older African American Normative Studies: Auditory Verbal Learning Test norms for African American elders. Clin. Neuropsychol. 19, 214–228, doi:10.1080/13854040590945300 (2005).

36 Jack, C. R., Jr. et al. Age, Sex, and APOE epsilon4 Effects on Memory, Brain Structure, and beta-Amyloid Across the Adult Life Span. JAMA Neurol 72, 511–519, doi:10.1001/jamaneurol.2014.4821 (2015).

37 Sliwinski, M. J. et al. Reliability and Validity of Ambulatory Cognitive Assessments. Assessment 25, 14–30, doi:10.1177/1073191116643164 (2016).

38 Ohman, F., Hassenstab, J., Berron, D., Scholl, M. & Papp, K. V. Current advances in digital cognitive assessment for preclinical Alzheimer’s disease. Alzheimers Dement (Amst) 13, e12217, doi:10.1002/dad2.12217 (2021).

39 Lohnas, L. J. & Kahana, M. J. Parametric effects of word frequency in memory for mixed frequency lists. J. Exp. Psychol. Learn. Mem. Cogn. 39, 1943–1946, doi:10.1037/a0033669 (2013).

40 Clark, J. M. & Paivio, A. Extensions of the Paivio, Yuille, and Madigan (1968) norms. Behav. Res. Methods Instrum. Comput. 36, 371–383, doi:10.3758/BF03195584 (2004).

41 Dolch, E. W. A basic sight vocabulary. The Elementary School Journal 36, 456–460, doi:10.1086/457353 (1936).

42 Therneau, T. M. A package for survival analysis in R, < https://CRAN.Rproject.org/package=survival> (2021).

43 Morrison, R. L. et al. A computerized, self-administered test of verbal episodic memory in elderly patients with mild cognitive impairment and healthy participants: A randomized, crossover, validation study. Alzheimers Dement (Amst) 10, 647–656, doi:10.1016/j.dadm.2018.08.010 (2018).

44 Belleville, S. et al. Neuropsychological Measures that Predict Progression from Mild Cognitive Impairment to Alzheimer’s type dementia in Older Adults: a Systematic Review and Meta-Analysis. Neuropsychol. Rev. 27, 328–353, doi:10.1007/s11065-017-9361-5 (2017).

45 Weissberger, G. H. et al. Diagnostic Accuracy of Memory Measures in Alzheimer’s Dementia and Mild Cognitive Impairment: a Systematic Review and Meta-Analysis. Neuropsychol. Rev. 27, 354–388, doi:10.1007/s11065-017-9360-6 (2017).

46 Atkinson, R. C. & Shiffrin, R. M. in The psychology of learning and motivation: II. (ed K. W. Spence & J. T. Spence) (Academic Press, 1968).

47 Gavett, B. E. & Horwitz, J. E. Immediate list recall as a measure of short-term episodic memory: insights from the serial position effect and item response theory. Arch. Clin. Neuropsychol. 27, 125–135, doi:10.1093/arclin/acr104 (2012).

48 Greene, J. D., Baddeley, A. D. & Hodges, J. R. Analysis of the episodic memory deficit in early Alzheimer’s disease: evidence from the doors and people test. Neuropsychologia 34, 537–551, doi:10.1016/0028-3932(95)00151-4 (1996).

49 Cunha, C., Guerreiro, M., de Mendonca, A., Oliveira, P. E. & Santana, I. Serial position effects in Alzheimer’s disease, mild cognitive impairment, and normal aging: predictive value for conversion to dementia. J. Clin. Exp. Neuropsychol. 34, 841–852, doi:10.1080/13803395.2012.689814 (2012).

50 Grober, E. & Kawas, C. Learning and retention in preclinical and early Alzheimer’s disease. Psychol. Aging 12, 183–188, doi:10.1037//0882-7974.12.1.183 (1997).

51 Stamate, A., Logie, R. H., Baddeley, A. D. & Della Sala, S. Forgetting in Alzheimer’s disease: Is it fast? Is it affected by repeated retrieval? Neuropsychologia 138, 107351, doi:10.1016/j.neuropsychologia.2020.107351 (2020).

52 Duke Han, S., Nguyen, C. P., Stricker, N. H. & Nation, D. A. Detectable Neuropsychological Differences in Early Preclinical Alzheimer’s Disease: A Meta-Analysis. Neuropsychol. Rev. 27, 305–325, doi:10.1007/s11065-017-9345-5 (2017).

53 Baker, J. E. et al. Cognitive impairment and decline in cognitively normal older adults with high amyloid-beta: A meta-analysis. Alzheimers Dement (Amst) 6, 108–121, doi:10.1016/j.dadm.2016.09.002 (2017).

54 Boyle, P. A. et al. To what degree is late life cognitive decline driven by age-related neuropathologies? Brain 144, 2166–2175, doi:10.1093/brain/awab092 (2021).

55 Harrington, K. D. et al. Estimates of age-related memory decline are inflated by unrecognized Alzheimer’s disease. Neurobiol. Aging 70, 170–179, doi:10.1016/j.neurobiolaging.2018.06.005 (2018).

56 Bos, I. et al. Amyloid-beta, Tau, and Cognition in Cognitively Normal Older Individuals: Examining the Necessity to Adjust for Biomarker Status in Normative Data. Front. Aging Neurosci. 10, 193, doi:10.3389/fnagi.2018.00193 (2018).

57 Alden, E. C. et al. Mayo normative studies: A conditional normative model for longitudinal change on the Auditory Verbal Learning Test and preliminary validation in preclinical Alzheimer’s disease. Alzheimers Dement (Amst) 14, e12325, doi:10.1002/dad2.12325 (2022).

58 Madero, E. N. et al. Environmental Distractions during Unsupervised Remote Digital Cognitive Assessment. The Journal of Prevention of Alzheimer’s Disease, 1–4, doi:10.14283/jpad.2021.9 (2021).

59 Lee, J. et al. The Overlap Index as a means of evaluating early tau-PET signal reliability. J. Nucl. Med., doi:10.2967/jnumed.121.263136 (2022).

60 Jack, C. R., Jr. et al. Associations of Amyloid, Tau, and Neurodegeneration Biomarker Profiles With Rates of Memory Decline Among Individuals Without Dementia. JAMA 321, 2316–2325, doi:10.1001/jama.2019.7437 (2019).

61 Papp, K. V. et al. The Computerized Cognitive Composite (C3) in an Alzheimer’s Disease Secondary Prevention Trial. J Prev Alzheimers Dis 8, 59–67, doi:10.14283/jpad.2020.38 (2021).

