## Supplemental Online Resources for "Stricker Learning Span criterion validity: a remote self-administered multi-device compatible digital word list memory measure shows similar ability to differentiate amyloid and tau PET-defined biomarker groups as in-person Auditory Verbal Learning Test"

### medRxiv

[1] Division of Neurocognitive Disorders, Department of Psychiatry and Psychology, Mayo Clinic, Rochester, Minnesota, USA
[2] Department of Information Technology, Mayo Clinic, Rochester, Minnesota, USA
[3] Division of Biomedical Statistics and Informatics, Department of Quantitative Health Sciences, Mayo Clinic, Rochester, Minnesota, USA
[4] Department of Radiology, Mayo Clinic, Rochester, Minnesota, USA
[5] Department of Neurology, Mayo Clinic, Rochester, Minnesota, USA
[6] Department of Epidemiology and Prevention, Wake Forest University School of Medicine, Winston-Salem, NC, USA

**This is supplemental online resources for a preprint / unpublished manuscript that is not yet peer reviewed.**

Copyright 2022 Mayo Foundation for Medical Education and Research.

**Supplemental Figure 1.** Full Pearson r correlation matrix for associations between AVLT and SLS variables.

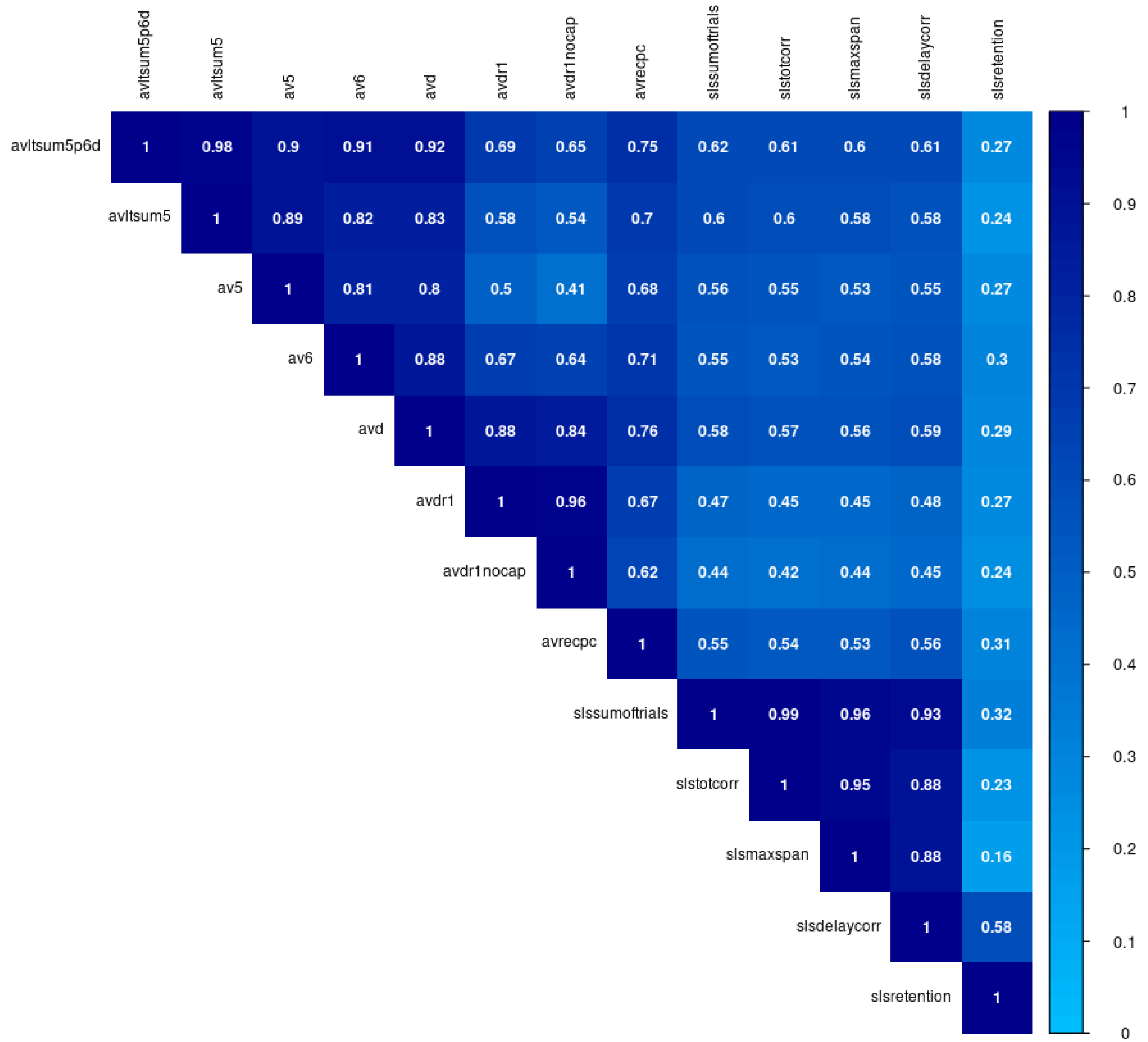


*Note*. All correlations are significant ($p$'s <0.001). AVLT = Auditory Verbal Learning Test; SLS = Stricker Learning Span;

avltsum5p6d = AVLT Sum of Trials (AVLT 1-5 total + Trial 6 + 30-minute delay); avltsum5 = AVLT Trials 1-5 total correct; av5 = AVLT Trial 5; av6 = AVLT Trial 6 (Short Delay); avd = AVLT 30-minute delay; avdr1 = long-term percent retention (AVLT 30-minute delay / Trial 5 but capped at 100% as typically done in the Mayo Clinic Study of Aging database); avdr1nocap = long-term percent retention (AVLT 30-minute delay / Trial 5 but without a cap, as presented in the primary manuscript for this study); avrecpc = AVLT Recognition Percent Correct {[recognition hits+(15 – recognition false positive errors)]/30} x 100; slssumoftrials = SLS Sum of Trials = SLS 1-5 total + delay; slstotcorr = SLS 1-5 Total (sum of words correctly recognized across trials 1-5); slsmaxspan = SLS Max Span (maximum number of words recognized across any of the 5 learning trials); slsdelaycorr = SLS delay correct; slsretention = SLS Retention (slsdelaycorr / slsmaxspan). Figure used with permission of Mayo Foundation for Medical Education and Research; all rights reserved.

**Supplemental Figure 2.** Scatterplot of SLS sum of trials and AVLT sum of trials (r=0.62). Figure used with permission of Mayo Foundation for Medical Education and Research; all rights reserved.

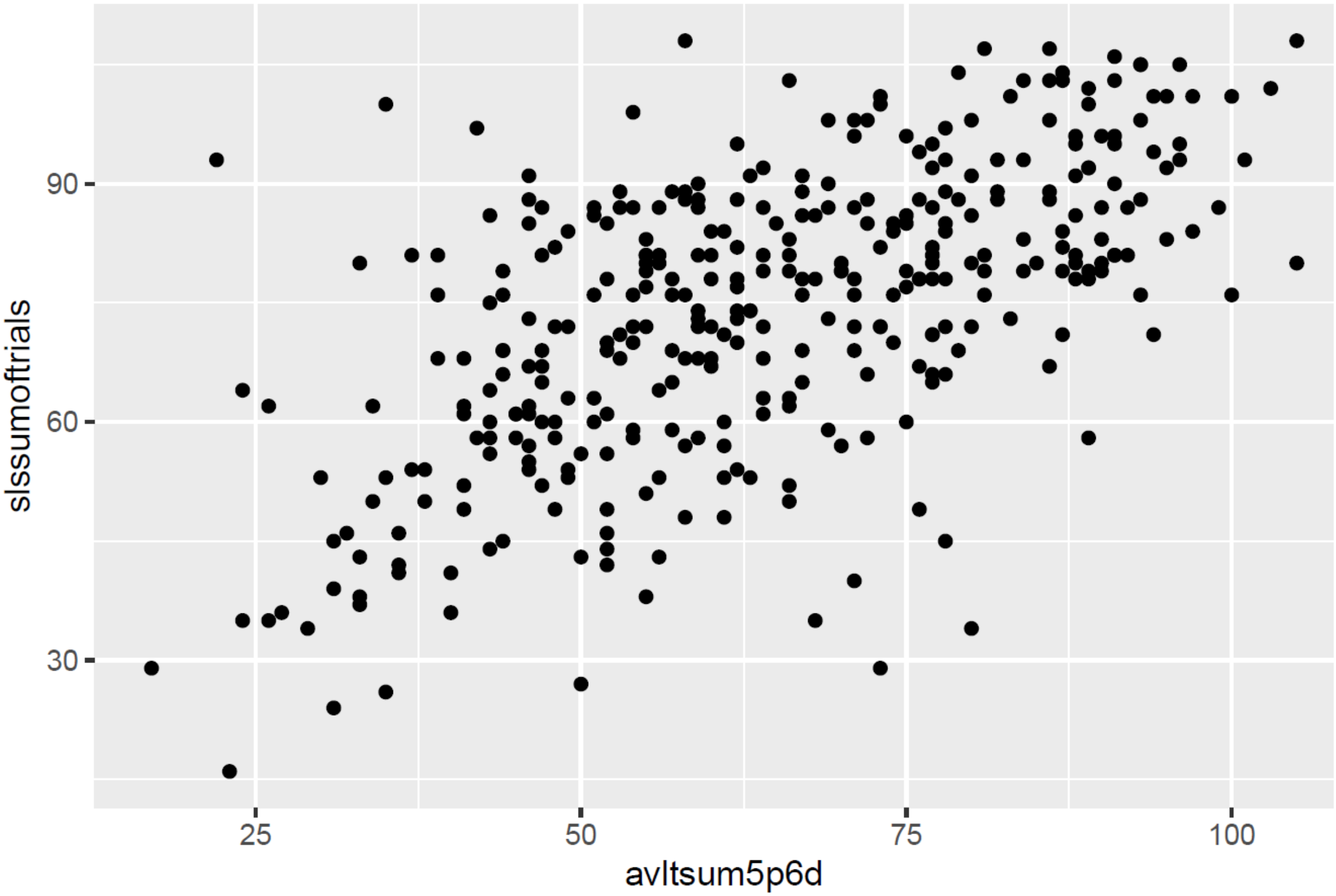

Supplemental Table 1. Additional memory test variable means (SDs) by biomarker subgroups.

| | A- vs A+ | | | | A-T- vs A+T+ | | | |
|---|---|---|---|---|---|---|---|---|
| | All participants | | CU participants only | | All participants | | CU participants only | |
| | A- (N=228) | A+ (N=125) | CU A- (N=215) | CU A+ (N=111) | A-T- (n=195) | A+T+ (n=55) | CU A-T- (n=185) | CU A+T+ (n=42) |
| SLS Trial 1 | 6.68 (1.25) | 6.27 (1.30) | 6.77 (1.19) | 6.36 (1.17) | 6.73 (1.24) | 6.04 (1.47) | 6.81 (1.19) | 6.21 (1.22) |
| AVLT Trial 1 | 6.71 (2.36) | 5.29 (2.03) | 6.86 (2.33) | 5.57 (1.96) | 6.72 (2.38) | 4.89 (1.71) | 6.84 (2.36) | 5.43 (1.52) |
| SLS Trial 2 | 10.04 (2.14) | 8.90 (2.54) | 10.20 (1.97) | 9.20 (2.35) | 10.07 (2.15) | 8.24 (2.52) | 10.21 (2.01) | 8.76 (2.20) |
| AVLT Trial 2 | 8.84 (2.65) | 7.42 (2.61) | 9.05 (2.56) | 7.79 (2.47) | 8.86 (2.66) | 6.98 (2.34) | 9.04 (2.59) | 7.71 (2.06) |
| SLS Trial 3 | 12.88 (3.04) | 11.47 (3.46) | 13.14 (2.84) | 11.89 (3.18) | 12.89 (3.02) | 10.76 (3.77) | 13.11 (2.87) | 11.52 (3.38) |
| AVLT Trial 3 | 10.18 (2.72) | 8.99 (2.92) | 10.38 (2.65) | 9.44 (2.70) | 10.24 (2.75) | 8.38 (2.81) | 10.42 (2.68) | 9.26 (2.40) |
| SLS Trial 4 | 15.72 (4.12) | 13.76 (4.80) | 16.04 (3.94) | 14.38 (4.47) | 15.90 (4.13) | 12.31 (4.83) | 16.15 (4.00) | 13.29 (4.48) |
| AVLT Trial 4 | 11.01 (2.71) | 9.93 (2.90) | 11.25 (2.55) | 10.41 (2.67) | 11.15 (2.66) | 9.15 (2.92) | 11.34 (2.54) | 10.07 (2.59) |
| SLS Trial 5 | 16.55 (4.06) | 14.63 (4.56) | 16.92 (3.81) | 15.28 (4.11) | 16.70 (4.05) | 13.33 (4.69) | 17.01 (3.85) | 14.48 (4.00) |
| AVLT Trial 5 | 11.50 (2.59) | 10.26 (2.86) | 11.71 (2.44) | 10.68 (2.70) | 11.63 (2.54) | 9.71 (2.67) | 11.80 (2.43) | 10.52 (2.33) |
| SLS 1-3 Total | 29.60 (5.64) | 26.64 (6.56) | 30.11 (5.16) | 27.45 (5.96) | 29.70 (5.60) | 25.04 (7.07) | 30.12 (5.19) | 26.50 (6.12) |
| AVLT 1-3 Total | 25.78 (7.11) | 22.17 (6.69) | 26.28 (6.94) | 22.80 (6.32) | 25.88 (7.16) | 21.21 (5.87) | 26.30 (7.03) | 22.40 (5.09) |
| SLS 1-4 Total | 45.32 (9.25) | 40.40 (10.96) | 46.15 (8.55) | 41.83 (10.00) | 45.59 (9.19) | 37.35 (11.57) | 46.27 (8.62) | 39.79 (10.27) |
| AVLT Ret. Alt. | 80.79 (20.76) | 70.73 (30.13) | 82.65 (18.95) | 77.29 (24.38) | 81.95 (19.38) | 63.36 (32.07) | 83.49 (17.67) | 77.14 (20.53) |
| Memory z [1] | 0.69 (1.01) | 0.74 (1.12) | 0.80 (0.91) | 0.87 (0.99) | 0.70 (1.00) | 0.45 (1.28) | 0.80 (0.91) | 0.71 (1.09) |
| LM I [1] | 27.54 (7.72) | 25.94 (7.88) | 28.27 (7.20) | 26.67 (7.38) | 27.70 (7.56) | 24.54 (8.03) | 28.31 (7.16) | 26.02 (6.99) |
| LM II [1] | 24.09 (9.19) | 22.08 (9.18) | 25.03 (8.44) | 22.95 (8.62) | 24.44 (8.89) | 19.77 (9.49) | 25.23 (8.25) | 21.43 (8.63) |
| VR I [1] | 30.98 (4.66) | 29.14 (5.30) | 31.40 (4.10) | 29.59 (4.93) | 31.22 (4.47) | 28.10 (5.74) | 31.56 (4.17) | 28.98 (5.08) |
| VR II [1] | 27.44 (7.17) | 24.14 (9.06) | 28.24 (6.24) | 25.16 (8.21) | 27.72 (6.96) | 22.77 (9.75) | 28.44 (6.05) | 24.79 (8.43) |

[1] Data unavailable for Alzheimer's Disease Research Center (ADRC) participants due to differences in test battery administered (n=8).

*Note*. AVLT = Auditory Verbal Learning Test; AVLT Ret. Alt. = AVLT Retention alternative with cap at 100% (typical calculation of retention in the Mayo Clinic Study of Aging (MCSA) data that differs from version presented in manuscript that does not use a cap); AVLT 1-3 Total = total correctly recalled trial 1 + trial 2 + trial 3; LM I = Wechsler Memory Scale-Revised Logical Memory I total; LM II = Wechsler Memory Scale-Revised Logical Memory II total; Memory z = Memory composite reflecting average (z) of three delayed recall scores including AVLT 30-minute delay, LM II and VR II (z-scores previously derived using CU participants 50 and older enrolled from 2004-2012, weighted by the age and sex distribution of the 2013 Olmsted county population [Rocca et al., 2021 DOI: 10.1136/bmjopen-2020-042633]); SLS = Stricker Learning Span; SLS 1-3 Total = total correct trial 1 + trial 2 + trial 3; SLS 1-4 Total = total correct trial 1 + trial 2 + trial 3 + trial 4. slst13totcorr (create new variable slsr1corr + slsr2corr + slsr3corr); VR I = Wechsler Memory Scale-Revised Visual Reproduction I total; VR II = Wechsler Memory Scale-Revised Visual Reproduction II total; total correct for fewer learning trials are presented to examine the potential impact of a shorter version of each test, such as what is provided in the NIH Toolbox Cognition app for the AVLT. Table used with permission of Mayo Foundation for Medical Education and Research; all rights reserved.

Supplemental Table 2. Unadjusted and adjusted Hedge's g effect sizes (95% CI) between biomarker subgroups for additional memory test variables.

| | A- vs A+ | | | | A-T- vs A+T+ | | | |
|---|---|---|---|---|---|---|---|---|
| | All A participants (N=353) | | CU A participants (N=326) | | All AT participants (N=259) | | CU AT participants (N=224) | |
| | Unadj g | Adj g | Unadj g | Adj g | Unadj g | Adj g | Unadj g | Adj g |
| SLS Trial 1 | -0.32* (-0.54, -0.1) | -0.17 (-0.38, 0.04) | -0.34* (-0.57, -0.11) | -0.20 (-0.42, 0.02) | -0.54* (-0.84, -0.23) | -0.30* (-0.55, -0.05) | -0.49* (-0.83, -0.16) | -0.26 (-0.52, 0.01) |
| AVLT Trial 1 | -0.63* (-0.85, -0.41) | -0.47* (-0.68, -0.26) | -0.58* (-0.82, -0.35) | -0.43* (-0.64, -0.21) | -0.81* (-1.12, -0.50) | -0.55* (-0.80, -0.30) | -0.63* (-0.97, -0.29) | -0.37* (-0.63, -0.11) |
| SLS Trial 2 | -0.50* (-0.72, -0.28) | -0.28* (-0.49, -0.07) | -0.47* (-0.71, -0.24) | -0.25* (-0.47, -0.03) | -0.82* (-1.13, -0.51) | -0.50* (-0.75, -0.25) | -0.70* (-1.05, -0.36) | -0.37* (-0.63, -0.11) |
| AVLT Trial 2 | -0.54* (-0.76, -0.32) | -0.31* (-0.52, -0.10) | -0.49* (-0.73, -0.26) | -0.27* (-0.48, -0.05) | -0.72* (-1.03, -0.42) | -0.41* (-0.66, -0.16) | -0.53* (-0.87, -0.19) | -0.23 (-0.49, 0.04) |
| SLS Trial 3 | -0.44* (-0.66, -0.22) | -0.19 (-0.4, 0.02) | -0.42* (-0.65, -0.19) | -0.17 (-0.39, 0.05) | -0.66* (-0.97, -0.36) | -0.32* (-0.58, -0.07) | -0.53* (-0.87, -0.19) | -0.18 (-0.45, 0.08) |
| AVLT Trial 3 | -0.42* (-0.65, -0.20) | -0.18 (-0.39, 0.03) | -0.35* (-0.58, -0.12) | -0.09 (-0.31, 0.13) | -0.67* (-0.97, -0.36) | -0.35* (-0.60, -0.10) | -0.44* (-0.78, -0.10) | -0.14 (-0.40, 0.12) |
| SLS Trial 4 | -0.45* (-0.67, -0.23) | -0.24* (-0.45, -0.03) | -0.40* (-0.63, -0.17) | -0.20 (-0.42, 0.02) | -0.83* (-1.14, -0.52) | -0.54* (-0.79, -0.29) | -0.70* (-1.04, -0.36) | -0.40* (-0.67, -0.14) |
| AVLT Trial 4 | -0.39* (-0.61, -0.17) | -0.13 (-0.34, 0.08) | -0.32 (-0.55, -0.09) | -0.05 (-0.27, 0.17) | -0.73* (-1.04, -0.43) | -0.40* (-0.65, -0.15) | -0.50* (-0.83, -0.16) | -0.18 (-0.44, 0.09) |
| SLS Trial 5 | -0.45* (-0.67, -0.23) | -0.23* (-0.44, -0.02) | -0.42* (-0.65, -0.19) | -0.17 (-0.39, 0.05) | -0.80* (-1.11, -0.49) | -0.48* (-0.73, -0.23) | -0.65* (-0.99, -0.31) | -0.30* (-0.57, -0.04) |
| AVLT Trial 5 | -0.46* (-0.68, -0.24) | -0.18 (-0.39, 0.03) | -0.41* (-0.64, -0.17) | -0.12 (-0.34, 0.1) | -0.74* (-1.05, -0.44) | -0.40* (-0.65, -0.15) | -0.53* (-0.87, -0.19) | -0.19 (-0.46, 0.07) |
| SLS 1-3 Total | -0.49* (-0.72, -0.27) | -0.25* (-0.46, -0.04) | -0.49* (-0.72, -0.26) | -0.24* (-0.46, -0.02) | -0.78* (-1.09, -0.47) | -0.43* (-0.68, -0.18) | -0.67* (-1.01, -0.33) | -0.30* (-0.57, -0.04) |
| AVLT 1-3 Total | -0.52* (-0.74, -0.29) | -0.26* (-0.48, -0.05) | -0.52* (-0.75, -0.28) | -0.28* (-0.5, -0.06) | -0.67* (-1.00, -0.35) | -0.32* (-0.58, -0.07) | -0.58* (-0.92, -0.24) | -0.27* (-0.53, 0.00) |
| SLS 1-4 Total | -0.50* (-0.72, -0.28) | -0.26* (-0.47, -0.05) | -0.48* (-0.71, -0.24) | -0.24* (-0.45, -0.02) | -0.84* (-1.15, -0.53) | -0.50* (-0.75, -0.25) | -0.72* (-1.06, -0.38) | -0.37* (-0.63, -0.11) |
| AVLT Ret. Alt. | -0.41* (-0.63, -0.19) | -0.20 (-0.41, 0.01) | -0.26* (-0.49, -0.03) | -0.06 (-0.28, 0.16) | -0.81* (-1.12, -0.51) | -0.53* (-0.78, -0.28) | -0.35* (-0.68, -0.01) | -0.15 (-0.41, 0.11) |
| Memory z [1] | 0.04 (-0.18, 0.26) | 0.01 (-0.20, 0.22) | 0.08* (-0.15, 0.31) | -0.02 (-0.23, 0.20) | -0.24 (-0.55, 0.08) | -0.25 (-0.51, 0.00) | -0.09 (-0.43, 0.24) | -0.20 (-0.46, 0.07) |
| LM I [1] | -0.21 (-0.43, 0.02) | 0.01 (-0.20, 0.22) | -0.22 (-0.45, 0.01) | -0.03 (-0.25, 0.19) | -0.41* (-0.73, -0.09) | -0.16 (-0.42, 0.09) | -0.32 (-0.66, 0.02) | -0.13 (-0.40, 0.13) |
| LM II [1] | -0.22 (-0.44, 0.00) | 0.02 (-0.20, 0.23) | -0.24* (-0.47, -0.01) | -0.04 (-0.26, 0.18) | -0.52* (-0.84, -0.20) | -0.24 (-0.49, 0.02) | -0.46* (-0.79, -0.12) | -0.23 (-0.49, 0.03) |
| VR I [1] | -0.38* (-0.60, -0.15) | -0.01 (-0.22, 0.20) | -0.41* (-0.64, -0.18) | -0.06 (-0.28, 0.16) | -0.65* (-0.98, -0.33) | -0.17 (-0.43, 0.08) | -0.59* (-0.93, -0.25) | -0.16 (-0.43, 0.10) |
| VR II [1] | -0.42 (-0.64, -0.19) | -0.08 (-0.29, 0.13) | -0.44* (-0.67, -0.21) | -0.10 (-0.32, 0.12) | -0.65* (-0.97, -0.33) | -0.22 (-0.48, 0.03) | -0.56* (-0.90, -0.22) | -0.16 (-0.43, 0.10) |

* $p < .05$

[1] Data unavailable for Alzheimer's Disease Research Center (ADRC) participants due to differences in test battery administered (n=8).
*Note*. AVLT = Auditory Verbal Learning Test; AVLT Ret. Alt. = AVLT Retention alternative with cap at 100% (typical calculation of retention in the Mayo Clinic Study of Aging (MCSA) data that differs from version presented in manuscript that does not use a cap); AVLT 1-3 Total = total correctly recalled trial 1 + trial 2 + trial 3; LM I = Wechsler Memory Scale-Revised Logical Memory I total; LM II = Wechsler Memory Scale-Revised Logical Memory II total; Memory z = Memory composite reflecting average (z) of three delayed recall scores including AVLT 30-minute delay, LM II and VR II; SLS = Stricker Learning Span; SLS 1-3 Total = total correct trial 1 + trial 2 + trial 3; SLS 1-4 Total = total correct trial 1 + trial 2 + trial 3 + trial 4. slst13totcorr (create new variable slsr1corr + slsr2corr + slsr3corr); VR I = Wechsler Memory Scale-Revised Visual Reproduction I total; VR II = Wechsler Memory Scale-Revised Visual Reproduction II total; total correct for fewer learning trials are presented to examine the potential impact of a shorter version of each test, such as what is provided in the NIH Toolbox Cognition app for the AVLT. Table used with permission of Mayo Foundation for Medical Education and Research; all rights reserved.
